# Enhancing the BOADICEA cancer risk prediction model to incorporate new data on *RAD51C, RAD51D, BARD1*, updates to tumour pathology and cancer incidences

**DOI:** 10.1101/2022.01.27.22269825

**Authors:** Andrew Lee, Nasim Mavaddat, Alex P. Cunningham, Tim Carver, Stephanie Archer, Fiona M. Walter, Marc Tischkowitz, Jonathan Roberts, Juliet Usher-Smith, Jacques Simard, Marjanka K. Schmidt, Peter Devilee, Vesna Zadnik, Hannes Jürgens, Emmanuelle Mouret-Fourme, Antoine De Pauw, Matti Rookus, Thea M. Mooij, Paul P.D. Pharoah, Douglas F. Easton, Antonis C. Antoniou

## Abstract

**Background:** BOADICEA for breast cancer and the epithelial ovarian cancer (EOC) models included in the CanRisk tool (www.canrisk.org) provide future cancer risks based on rare pathogenic variants in cancer-susceptibility genes, polygenic risk scores, breast-density, questionnaire-based risk factors and family history. Here, we extend the models to include the effects of pathogenic variants in recently established breast cancer and EOC susceptibility genes, up-to-date age-specific pathology distributions and continuous risk factors.

**Methods:** BOADICEA was extended to further incorporate the associations of pathogenic variants in *BARD1, RAD51C* and *RAD51D* with breast cancer risk. The EOC model was extended to include the association of *PALB2* pathogenic variants with EOC risk. Age-specific distributions of oestrogen-receptor-negative and triple-negative breast cancer status for pathogenic variant carriers in these genes and *CHEK2* and *ATM* were also incorporated. A novel method to include continuous risk factors was developed, exemplified by including adult-height as continuous.

**Results:** *BARD1, RAD51C* and *RAD51D* explain 0.31% of the breast cancer polygenic variance. When incorporated into the multifactorial model, 34-44% of these carriers would be reclassified to the near-population and 15-22% to the high-risk categories based on the UK NICE guidelines. Including height as continuous, increased the BC relative-risk variance from 0.002 to 0.010.

**Conclusions:** These extensions will allow for better personalised risks for *BARD1, RAD51C, RAD51D* and *PALB2* pathogenic variant carriers and more informed choices on screening, prevention, risk factor modification or other risk-reducing options.

## Introduction

Breast cancer (BC) and epithelial tubo-ovarian cancer (EOC) are two of the most common cancers in females^1 2^. Through mammography or other methods, screening for BC can reduce mortality, and organised screening is available in most developed countries^3^. For EOC, no effective screening exists, but the disease can be prevented by salpingo-oophorectomy. However, these preventative options are associated with adverse effects. Therefore, identifying those at increased risk may help to target screening and preventative options to those most likely to benefit^4^. Both BC and EOC risks are multifactorial diseases, with family history of cancer (FH), genetic factors and lifestyle, hormonal and reproductive risk factors (RF) all contributing to risk^5-7^.

Previously we developed the BOADICEA (Breast and Ovarian Analysis of Disease Incidence and Carrier Estimation Algorithm) model for BC risk prediction and for the likelihood of carrying pathogenic variants (PVs) in BC susceptibility genes. BOADICEA v5 incorporates the effects of PVs in five BC susceptibility genes (*BRCA1, BRCA2, PALB2, CHEK2* and *ATM*), the effects of known common genetic variants summarised as a polygenic risk score (PRS), and a polygenic component that accounts for any residual familial aggregation^8 9^. We also developed a similar EOC model (Ovarian Cancer Model v1) that considers the effects of PVs in *BRCA1, BRCA2, RAD51D, RAD51C* and *BRIP1* on EOC together with a polygenic component^10 11^. BOADICEA includes mammographic density and both models incorporate the effects of known lifestyle, hormonal, reproductive and anthropometric RFs. In addition, the models incorporate breast tumour heterogeneity by considering the distributions of tumour oestrogen receptor (ER) and triple-negative (TN) (ER, progesterone receptor and human epidermal growth factor receptor 2 negative) status for *BRCA1* and *BRCA2* PV carriers and the general population^12 13^. Both models are freely available to healthcare professionals via the CanRisk webtool (www.canrisk.org), and are widely used by healthcare professionals^14^.

Recently, large population-based and family-based targeted sequencing studies have established that PVs in *RAD51C, RAD51D* and *BARD1* are associated with BC risk^15 16^ and that PVs in *PALB2* are associated with EOC risk^17 18^. In addition, analysis of the tumour characteristics in the BRIDGES study has provided age-specific estimates of the distributions of tumour characteristics for PV carriers in all established susceptibility genes^19^.

A further limitation of the previous models is that all epidemiological RFs are treated as categorical. However, some RFs (e.g., height, body mass index (BMI) mammographic density) are intrinsically continuous, and discretisation results in a loss of information, reducing their predictive ability.

Here we extend both models to explicitly model the effects of PVs in the recently established BC and EOC susceptibility genes and incorporate up-to-date age-specific pathology distributions. We present a methodological framework for incorporating continuous RFs into the model, and we demonstrate this by including height as a continuous variable. Finally, we describe updates to the population reference cancer incidences used in the models by incorporating more up-to-date incidences, incidences for additional countries and refining the derivation of birth-cohort specific incidences for inclusion in the models that address sparsity in the population incidence data.

## Methods

### Rare Moderate-Risk Pathogenic Variants

Both BOADICEA and the EOC Model model cancer incidence as an explicit function of PVs in known high- and moderate-penetrance susceptibility genes (major genes) together with a polygenic component^9-12 20-22^. By using an explicit genetic model, they can account for both genetic testing and detailed FH. BOADICEA includes the genes *BRCA1, BRCA2, PALB2, CHEK2* and *ATM*, with dominance in that order, along with a BC susceptibility polygenic component. The EOC Model includes the genes *BRCA1, BRCA2, RAD51D, RAD51C* and *BRIP1*, with dominance in that order, along with an EOC susceptibility polygenic component. Details of the underlying model are included in the supplementary material. The values of the parameters for the original models were determined by complex segregation analysis^9 10^. However, this was not possible for the extended versions since no sufficiently large data set containing all the model features was available. Instead, we adopted a synthetic approach^23^, in which additional model parameters are taken from large-scale external studies^8 11 12 21^.

Here, BOADICEA was extended to explicitly model the effects of PVs in *BARD1, RAD51C* and *RAD51D* (eight BC susceptibility genes), while the EOC model was extended to include *PALB2* (six EOC susceptibility genes). In both models, the effects of PVs were included as major genes and are parameterised by their allele frequency in the general population and their age-specific relative risks (RR). The BC RR for carriers of PV in *BARD1* was taken from the BRIDGES study^15^, while those for *RAD51C* and *RAD51D* were the meta-analysed values from Dorling et al^15^ and Yang et al^24^. The EOC RR for *PALB2* PV carriers was taken from Yang et al^17^. Updated RRs for carriers of PV in *ATM*, along with PV carrier frequencies for *PALB2, CHEK2, ATM, BARD1, RAD51D, RAD51C* and *BRIP1* and screening test sensitivities for all genes were derived from Dorling et al^15^. We used the BRIDGES study to derive these frequency estimates as it is a very large population-based dataset that includes targeted sequencing data. Frequencies were based on the control frequencies in European populations, adjusted for the assumed sensitivity of the sequencing and the fact that large rearrangements were not detected (supplementary material). The default sensitivities were then calculated, assuming that clinical genetic testing will detect all known pathogenic mutations except for large rearrangements (except *BRCA1* and *BRCA2*, where testing for large rearrangements is routinely done). All model parameters for PVs are given in Table 1.

**Table 1.**
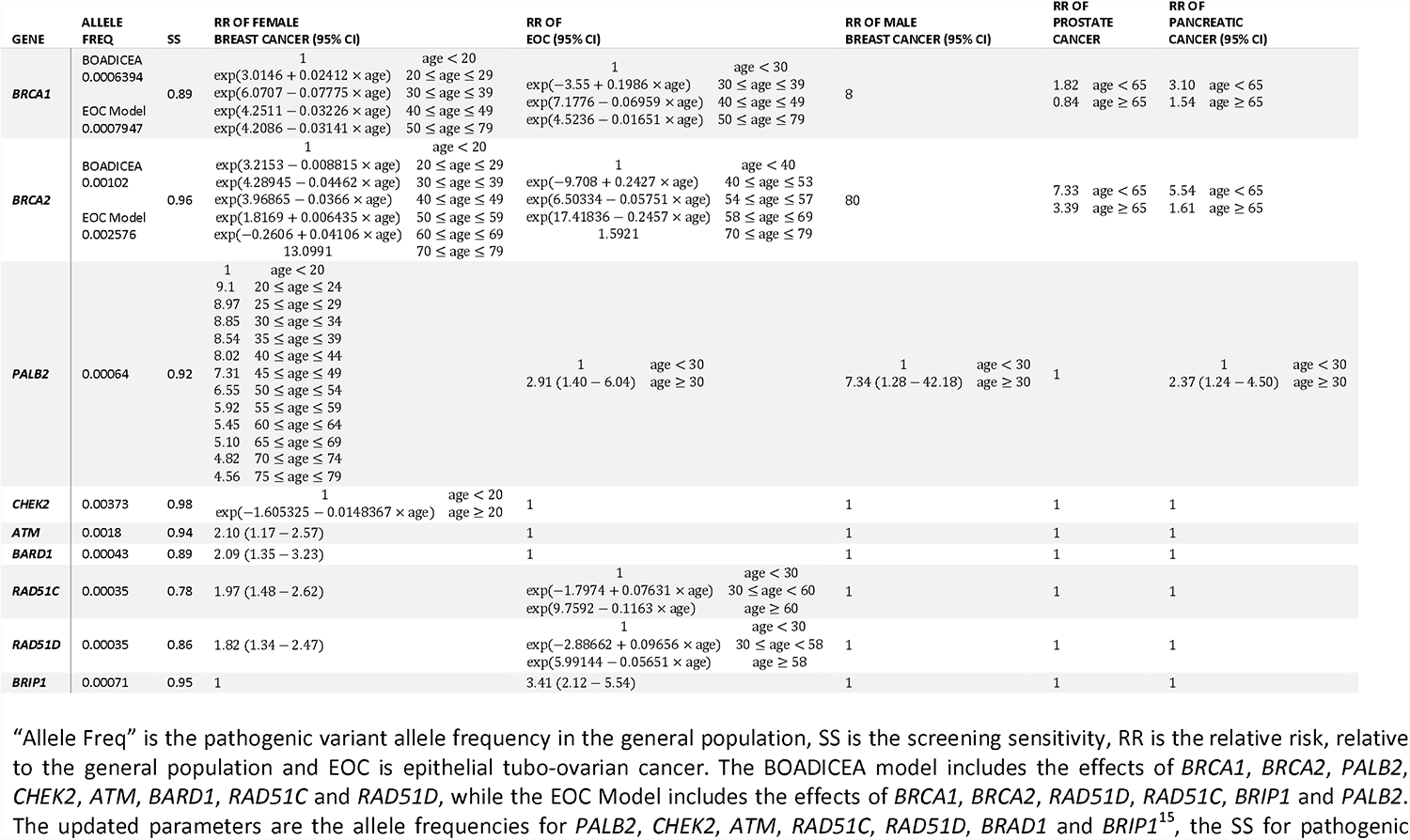
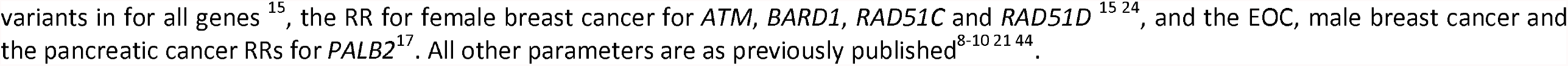
The parameters used to include the effects of rare high- and intermediate-risk pathogenic variants in the models.

| GENE | ALLELE<br>FREQ | SS | RR OF FEMALE<br>BREAST CANCER (95% CI) | RR OF<br>EOC (95% CI) | RR OF MALE<br>BREAST CANCER (95% CI) | RR OF<br>PROSTATE<br>CANCER | RR OF<br>PANCREATIC<br>CANCER (95% CI) |
| --- | --- | --- | --- | --- | --- | --- | --- |
| **BRCA1** | BOADICEA<br>0.0006394 | 0.89 | 1<br>age < 20<br>exp(3.0146 + 0.02412 × age)<br>20 ≤ age ≤ 29<br>exp(6.0707 – 0.07775 × age)<br>30 ≤ age ≤ 39<br>exp(4.2511 – 0.03226 × age)<br>40 ≤ age ≤ 49<br>exp(4.2086 – 0.03141 × age)<br>50 ≤ age ≤ 79 | 1<br>age < 30<br>exp(–3.55 + 0.1986 × age)<br>30 ≤ age ≤ 39<br>exp(7.1776 – 0.06959 × age)<br>40 ≤ age ≤ 49<br>exp(4.5236 – 0.01651 × age)<br>50 ≤ age ≤ 79 | 8 | 1.82<br>age < 65<br>0.84<br>age ≥ 65 | 3.10<br>age < 65<br>1.54<br>age ≥ 65 |
|  | EOC Model<br>0.0007947 |  |  |  |  |  |  |
| **BRCA2** | BOADICEA<br>0.00102 | 0.96 | 1<br>age < 20<br>exp(3.2153 – 0.008815 × age)<br>20 ≤ age ≤ 29<br>exp(4.28945 – 0.04462 × age)<br>30 ≤ age ≤ 39<br>exp(3.96865 – 0.0366 × age)<br>40 ≤ age ≤ 49<br>exp(1.8169 + 0.006435 × age)<br>50 ≤ age ≤ 59<br>exp(–0.2606 + 0.04106 × age)<br>60 ≤ age ≤ 69<br>13.0991<br>70 ≤ age ≤ 79 | 1<br>age < 40<br>exp(–9.708 + 0.2427 × age)<br>40 ≤ age ≤ 53<br>exp(6.50334 – 0.05751 × age)<br>54 ≤ age ≤ 57<br>exp(17.41836 – 0.2457 × age)<br>58 ≤ age ≤ 69<br>1.5921<br>70 ≤ age ≤ 79 | 80 | 7.33<br>age < 65<br>3.39<br>age ≥ 65 | 5.54<br>age < 65<br>1.61<br>age ≥ 65 |
|  | EOC Model<br>0.002576 |  |  |  |  |  |  |
| **PALB2** | 0.00064 | 0.92 | 1<br>age < 20<br>9.1<br>20 ≤ age ≤ 24<br>8.97<br>25 ≤ age ≤ 29<br>8.85<br>30 ≤ age ≤ 34<br>8.54<br>35 ≤ age ≤ 39<br>8.02<br>40 ≤ age ≤ 44<br>7.31<br>45 ≤ age ≤ 49<br>6.55<br>50 ≤ age ≤ 54<br>5.92<br>55 ≤ age ≤ 59<br>5.45<br>60 ≤ age ≤ 64<br>5.10<br>65 ≤ age ≤ 69<br>4.82<br>70 ≤ age ≤ 74<br>4.56<br>75 ≤ age ≤ 79 | 1<br>age < 30<br>2.91 (1.40 – 6.04)<br>age ≥ 30 | 1<br>age < 30<br>7.34 (1.28 – 42.18)<br>age ≥ 30 | 1 | 1<br>age < 30<br>2.37 (1.24 – 4.50)<br>age ≥ 30 |
| **CHEK2** | 0.00373 | 0.98 | 1<br>age < 20<br>exp(–1.605325 – 0.0148367 × age)<br>age ≥ 20 | 1 | 1 | 1 | 1 |
| **ATM** | 0.0018 | 0.94 | 2.10 (1.17 – 2.57) | 1 | 1 | 1 | 1 |
| **BARD1** | 0.00043 | 0.89 | 2.09 (1.35 – 3.23) | 1 | 1 | 1 | 1 |
| **RAD51C** | 0.00035 | 0.78 | 1.97 (1.48 – 2.62) | 1<br>age < 30<br>exp(–1.7974 + 0.07631 × age)<br>30 ≤ age < 60<br>exp(9.7592 – 0.1163 × age)<br>age ≥ 60 | 1 | 1 | 1 |
| **RAD51D** | 0.00035 | 0.86 | 1.82 (1.34 – 2.47) | 1<br>age < 30<br>exp(–2.88662 + 0.09656 × age)<br>30 ≤ age < 58<br>exp(5.99144 – 0.05651 × age)<br>age ≥ 58 | 1 | 1 | 1 |
| **BRIP1** | 0.00071 | 0.95 | 1 | 3.41 (2.12 – 5.54) | 1 | 1 | 1 |

“Allele Freq” is the pathogenic variant allele frequency in the general population, SS is the screening sensitivity, RR is the relative risk, relative to the general population and EOC is epithelial tubo-ovarian cancer. The BOADICEA model includes the effects of *BRCA1*, *BRCA2*, *PALB2*, *CHEK2*, *ATM*, *BARD1*, *RAD51C* and *RAD51D*, while the EOC Model includes the effects of *BRCA1*, *BRCA2*, *RAD51D*, *RAD51C*, *BRIP1* and *PALB2*. The updated parameters are the allele frequencies for *PALB2*, *CHEK2*, *ATM*, *RAD51C*, *RAD51D*, *BRAD1* and *BRIP1*<sup>15</sup>, the SS for pathogenic
variants in for all genes<sup>15</sup>, the RR for female breast cancer for *ATM*, *BARD1*, *RAD51C* and *RAD51D*<sup>15 24</sup>, and the EOC, male breast cancer and the pancreatic cancer RRs for *PALB2*<sup>17</sup>. All other parameters are as previously published<sup>8-10 21 44</sup>.

As the polygenic component captures all residual familial aggregation not explained by the major genes, the previous models implicitly included the contributions of PVs in the new genes (i.e., *BARD1, RAD51C* and *RAD51D* for BOADICEA and *PALB2* for the EOC Model). Therefore, to avoid double counting their contribution, it was necessary to remove their contribution from the polygenic component by adjusting the log-RR per standard deviation of the polygenic component such that the total variance of the polygenic component and the new genes is the same as that of the polygenic component of the previous model^21^ (supplementary material).

The association between *PALB2* PVs and EOC was also included in the BOADICEA model, and the associations with male BC and PaC have been included in both models^17^.

The effects of PV in the new BC susceptibility genes on risk prediction were assessed by considering the risk categories described in the National Institute for Health and Care Excellence (NICE) familial BC guidelines^25^. For lifetime risk (age 20 to 80 years), three categories are defined: 1) near-population-risk, for risks less than 17%, 2) moderate-risk, for risks in the range [17%, 30%) and 3) high-risk, for risks of 30% or greater. Reclassification was considered based on questionnaire-based RFs (QRF) (RFs other than mammographic density), mammographic density (MD, based on the BI-RADS system) and a polygenic risk score (PRS). For BC, the PRS was taken to be the Breast Cancer Association Consortium 313 variant PRS, which accounts for 20% of the overall polygenic variance^8 26^. For EOC, we defined three risk categories based on lifetime risk^27 28^: 1) near-population-risk, for risks of less than 5%, 2) moderate-risk, for risks in the range [5%, 10%) and 3) high-risk, for risks of 10% or greater, and reclassification was considered based on RFs and a PRS. For EOC, the PRS was taken as the Ovarian Cancer Association Consortium 36 variant PRS, which accounts for 5% of the overall polygenic variance^11 29^.

### Updates to Tumour Pathology

Both models incorporate data on BC tumour pathology, specifically ER and TN. The distribution of pathology for affected carriers of PVs differs substantially from that in non-carriers for several genes, so that pathology data can affect the carrier probabilities and hence cancer risks^11 12^. In BOADICEA and the EOC model, breast tumours are classified into five groups based on ER and TN status: ER unknown, ER-positive, ER-negative/TN unknown, ER-negative/not TN, and TN. Previously, the models achieved this using age-dependent distributions in the general population and *BRCA1* and *BRCA2* PV carriers and an age-independent distribution for *CHEK2* PV carriers^12 21^. Due to a lack of data, the tumour ER distribution for carriers of PV in other genes was assumed to be the same as the general population. Here, the models have been updated to incorporate age-dependent ER and TN tumour distributions for carriers of PVs in the BC susceptibility genes *PALB2, CHEK2, ATM, BARD1, RAD51C* and *RAD51D*, using data from BRIDGES^19^.

### Continuous Risk Factors

The previous versions of the models included reproductive, lifestyle, hormonal and anthropometric RFs^8 11^. One limitation of these models was that the RFs needed to be coded as categorical variables. Some RFs are naturally continuous, requiring prior discretisation to a finite number of categories, resulting in some loss of information and reduction in risk discrimination. Here, the methodology was extended to allow the inclusion of continuous risk factors.

The key challenge is to calculate the baseline incidences *λ*_0_ (*t*) in equation (1) (supplementary material) from the population incidence and the risk factor distributions. The baseline incidences are calculated sequentially for each age *t* (considered discrete) using the values at age *t*− 1, starting from age 0, requiring the evolution with age of the probability distribution of those who are disease-free ^30^. For discrete factors/genes, this involves summing over all possible categories/genotypes, but for continuous factors/genes, it would involve integrating over all possible values. In principle, these integrals could be computed (either analytically or numerically). However, at each age, the number of terms in the integrand increases by a factor of 2, so by age 80, there are >10^24^ terms, with evaluation becomes impracticable. Alternatively, the risk factor could be discretised into a very large number of categories. This would give a very close approximation to the continuous distribution, but (particularly once multiple risk factors are considered, as here) the large number of categories would also make the calculations impractical. Instead, we propose an alternate approach in which the continuous factors are discretised with categories adapted according to the observed RF. The approach is as follows:

1. Firstly, discretise the range of possible risk factor values into a finite number (*n*) of bins and calculate the probability mass and RR for each bin from the probability density and RR function for the continuous RF. This part is identical to the standard approach for discretising risk factors, used in the existing models^8^. For a risk factor, *x*, with probability density *P*(*x*) and relative risk *RR*(*x*) the probability mass for bin *i* with range [*l*_*i*_, *u*_*i*_] is:

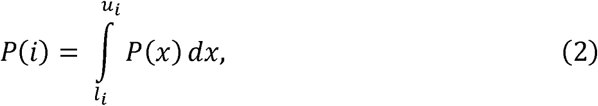

and the corresponding RR is

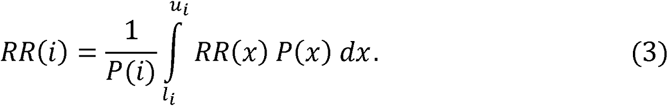
2. Create an additional (*n*+ 1)^*th*^ bin based on the individual’ s measured risk factor value that has an infinitesimal width. The RR for this bin is taken as the RR at the measured value, and it has zero mass. As this bin is infinitesimal, its overlap with the other bins is zero, so there is no double-counting.

This procedure creates a categorical risk factor with *n*+ 1 categories, where the individual is assigned to the (*n*+ 1)^*th*^category defined in step 2. This allows the exact value of the risk for the individual to be used, while the number of categories required to compute the baseline rates is fixed, limiting the computation time.

The accuracy of the approximation in the procedure relies on the assumption that the range of values within each bin have similar RRs, which should be reflected in the choice of discretisation scheme and the number of bins *n*. These choices will depend on the shape of the distribution and the RR function.

The above procedure can be applied to any risk factor distribution or RR function. However, the process assumes that an individual’ s position within the distribution is fixed with respect to age, although the value of the risk factor and RR may vary with age. Here, the method was applied to height.

### Updates to Population Incidences

The baseline incidences in equation (1) are birth-year and country specific as a consequence of using birth-year and country-specific population incidences in the constraining process. We refined the derivation of cohort-specific population incidences to account for variability in the incidences due to small numbers. In addition, we have updated existing incidences in the model to include more recent calendar periods and adapted the model to utilise cancer incidences from four new populations: the Netherlands, France, Slovenia and Estonia. Details are included in the supplementary material.

## Results

### Rare Moderate-Risk Pathogenic Variants

Table 1 summarises the models’ genetic parameter estimates, including those for the new genes. The estimated cumulative age-specific BC risks for *BARD1, RAD51C* and *RAD51D* PV carriers in BOADICEA and EOC risks for *PALB2* carriers, assuming the UK incidences applicable to those born in the 1980s, are shown in figure 1. The estimated average lifetime BC risks for PV carriers are 24%, 22% and 21% for *BARD1, RAD51C* and *RAD51D* PV carriers, respectively. The estimated lifetime EOC risk for *PALB2* carriers is 5.0%. Based on the assumed allele frequencies, 0.22% of the population carry PV in the genes *BARD1, RAD51C* or *RAD51D*, and these explain on average 0.31% of the female BC polygenic variance (averaged over all ages and cohorts, weighted by the age- and cohort-specific BC incidences). Approximately 0.13% of the population carry PVs in *PALB2*, explaining 0.16% of the EOC polygenic variance and 2.5% of the male BC polygenic variance.

**Figure 1:**
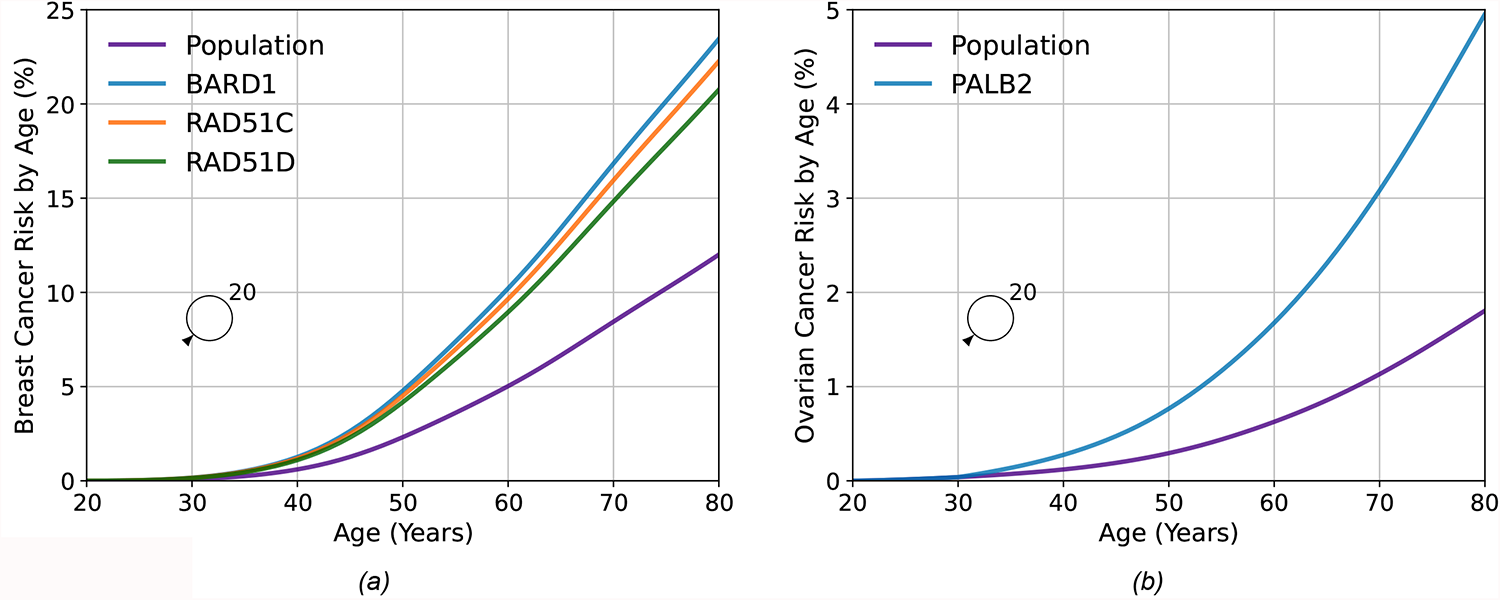
Predicted risk by age Vs age for a female born in 1985 with an unknown family history based on pathogenic variant carrier status for the new genes in the model. Figure (a) shows the breast cancer risk for carriers of pathogenic variants in *BARD1, RAD51C* and *RAD51D* along with the population risk. Figure (b) shows the ovarian cancer risk for carriers of pathogenic variants in *PALB2* along with the population risk. Predictions are based on UK cancer incidences.

Figure 2 (a)-(f) and Supplementary Table s1 show the distributions of lifetime BC risks for carriers of PVs in *BARD1, RAD51C* and *RAD51D* for a female with unknown FH and a female whose mother is affected at age 50 based on PV carrier status alone and including QRF, MD and a PRS. Based solely on PV carrier status, all females with unknown FH would be classified as at moderate risk. When information on QRF, MD or PRS is known, there is significant reclassification to near-population and high-risk categories, which is greatest when all factors are used in combination. For example, based on lifetime BC risks and using the full multifactorial model incorporating QRF, MD and a PRS, 33.9% of *BARD1* PV carriers with unknown FH would be reclassified from moderate-risk to near-population-risk, and 21.9% would be reclassified to high-risk (Supplementary Table s1). Similarly, BARD1 PV carriers with an affected first-degree relative would be considered high-risk (risk of 33.7% by age 80) based on family history and PV status alone. Incorporating the other risk factors would reclassify 12% as near-population risk and 40.2% as moderate-risk (Supplementary Table s1).

**Figure 2.**
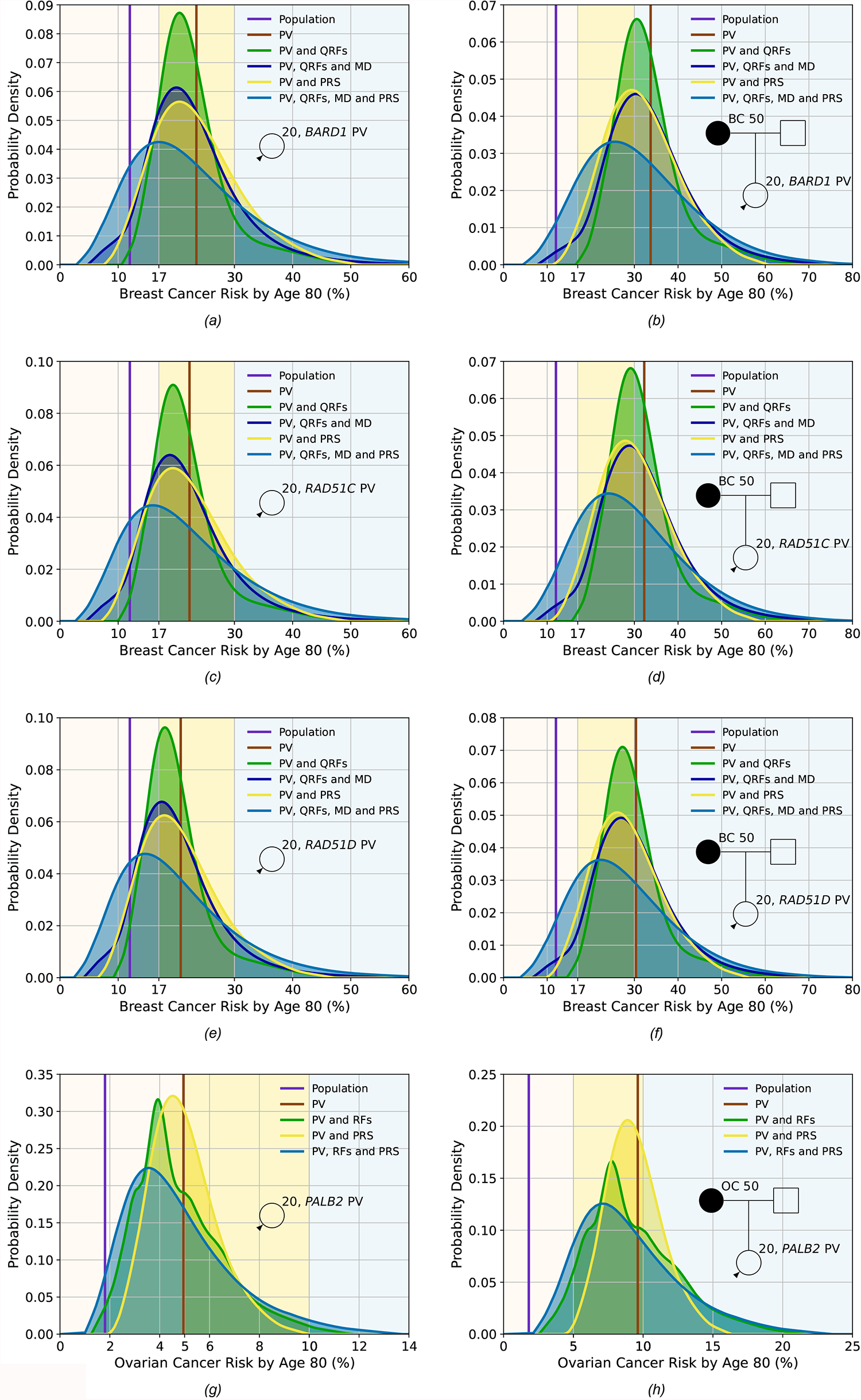
Predicted lifetime cancer risks (from age 20 to 80 years) for a female born in 1985 with a pathogenic variant in *BARD1* (breast cancer risk), *RAD51C* (breast cancer risk), *RAD51D* (breast cancer risk), and *PALB2* (ovarian cancer risk) on the basis of the different predictors of risk (pathogenic variant status (PV), questionnaire-based risk factors (QRFs), mammographic density (MD), and PRS). All figures show the probability density against the absolute risk. Figures (a), (c), (e) and (g) show risks for a female with unknown family history, while Figures (b), (d), (f) and (h) show risks where the individual’ s mother has had cancer at age 50. The backgrounds of the graphs are shaded to indicate the risk categories. For breast cancer, these are the categories defined by the National Institute for Health and Care Excellence familial breast cancer guidelines^25^: 1) near-population risk shaded in pink (<17%), 2) moderate risk shaded in yellow (≥17% and <30%) and 3) high risk shaded in blue (≥30%). For ovarian cancer, the categories are: 1) near-population risk shaded in pink (<5%), 2) moderate risk shaded in yellow (≥5% and <10%) and 3) high risk shaded in blue (≥10%). Predictions were based on UK cancer incidences. The line labelled population denotes the average population risk in the absence of knowledge of family history, pathogenic variant status, RFs or a PRS.

Figure 2 (g) and (h) and Supplementary Table s2 show the distribution of lifetime EOC risks for carriers of PVs in *PALB2* for a female with unknown FH and a female whose mother is affected at age 50, as a function of the RFs and PRS. For a *PALB2* carrier with unknown FH, when the RFs and PRS are considered jointly, 62.4% are classified as near-population-risk, 34.9% as moderate-risk, and 2.7% as high-risk. The corresponding proportions with an affected mother are 11.2%, 55.8%, and 33%, respectively. However, even among *PALB2* carriers with an affected mother, 97.5% will have risks of less than 3% by age 50 (Supplementary Table s2).

### Tumour Pathology

Figure 3 and Supplementary Tables s3 and s4 show the age-specific distributions of ER-negative tumours and TN tumours among ER-negative tumours used in the models for *PALB2, ATM, CHEK2, BARD1, RAD51C* and *RAD51D* PV carriers based on the BRIDGES data^19^. *BARD1, RAD51C* and *RAD51D* PV carriers predominantly develop ER-negative BCs, and the proportions decrease with increasing age. On the other hand, *CHEK2* and *ATM* carriers primarily develop ER-positive BCs, and the proportion of ER-positive tumours increases with age. Among those with ER-negative tumours, most tumours are TN for PV carriers in all genes, except *CHEK2* carriers, in whom the majority are ER-negative but not TN.

**Figure 3.**
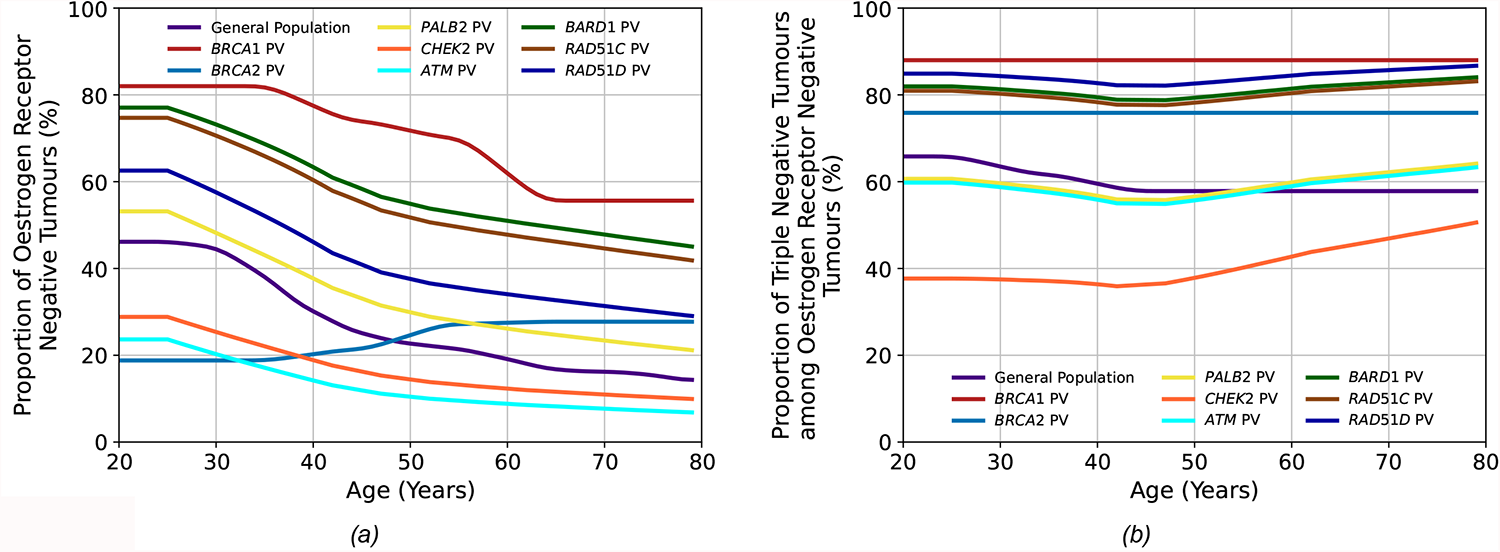
The tumour pathology proportions in the general population and among carriers of pathogenic variants (PV) in the breast cancer (BC) susceptibility genes included in the BOADICEA model. Figure (a) shows the proportion of oestrogen receptor-negative (ER-) tumours among all tumours, and Figure (b) shows the proportion of triple-negative (TN) (ER-, progesterone receptor-negative and human epidermal growth factor receptor 2) tumours among ER-tumours. The general population, *BRCA1* PV and *BRCA2* PV values are the same as previously used in the model^12^, while those for the other genes are updated using recent BRIDGES data^19^.

Using the updated age- and gene-specific ER-negative and TN tumour status distributions resulted in differences in the predicted carrier probabilities by different tumour pathology and age (Figure 4). For *ATM*, the carrier probabilities for ER-negative tumours are reduced relative to previous estimates, reflecting the stronger association with ER-positive disease. Carrier probabilities for *CHEK2* now show a decline with age for ER-negative tumours (previously, this was only predicted for ER-positive disease). The carrier probabilities for *PALB2* remain similar to previous estimates. For the new genes *BARD1, RAD51C* and *RAD51D*, the carrier probabilities are, as expected, higher for ER-negative and TN disease, but there is little variation by age.

**Figure 4:**
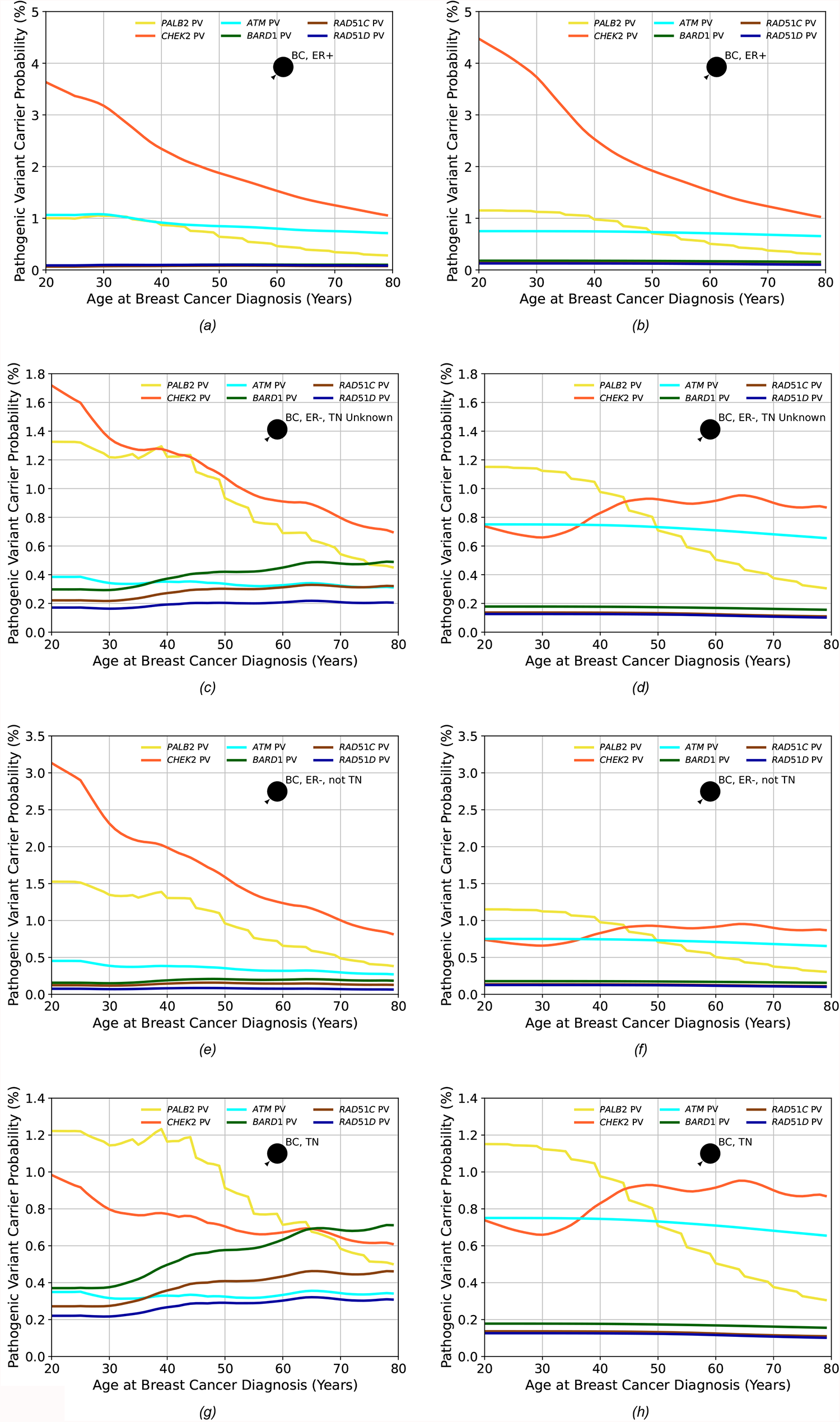
The probabilities of carrying a pathogenic variant estimated by BOADICEA in the genes *PALB2, CHEK2, ATM, BARD1, RAD51C* and *RAD51D* for an affected female born in 1985 as a function of her age at diagnosis based on different tumour pathology. Figures (a), (c), (e) and (g) show the probabilities based on the updated proportions, while figures (b), (d), (f) and (h) are based on the previous proportions and where proportions for *BARD1, RAD51C* and *RAD51D*, which were not in the previous model, are assumed to be the same as in the general population. In figures (a) and (b), the female has had an oestrogen receptor-positive (ER+) tumour; in figures (c) and (d), the female has had an oestrogen receptor-negative (ER-) tumour, but the triple-negative (TN) status is unknown; in figures (e) and (f), the female has had an ER- tumour that is not TN and in figures (g) and (h), the female has had a TN tumour. Predictions are based on UK cancer incidences.

### Continuous Risk Factors

As previously, adult female height was assumed to be normally distributed with mean 162.81cm and standard deviation 6.452cm, and be associated a log-RR per standard deviation, for both breast and ovarian cancer, of 0.10130^8 11^. We therefore discretised the normal distribution such that the probability masses of the bins were given by a binomial distribution 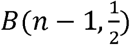, giving sufficient discretisation to adequately capture the tails of the distribution. We examined the relative discretisation error of the predicted lifetime risk as a function of the number of bins (Figure 5 e and f) and chose *n* = 5, as the lowest number of bins such that the root-mean-square relative error was less than 10^−4^. Compared to the discrete (five-level) RF, the variance of the RR of both BC and EOC increased from 0.002 to 0.010 when height was included as a continuous RF. The effects on predicted lifetime risks are shown in Figure 5 (a)-(d). Under the continuous implementation here, the lifetime BC risk varied from 9.7% for the first percentile to 14.6% for the 99^th^, whereas under the previous discrete distribution, the risks range from 10.1% to 14.2%.

**Figure 5.**
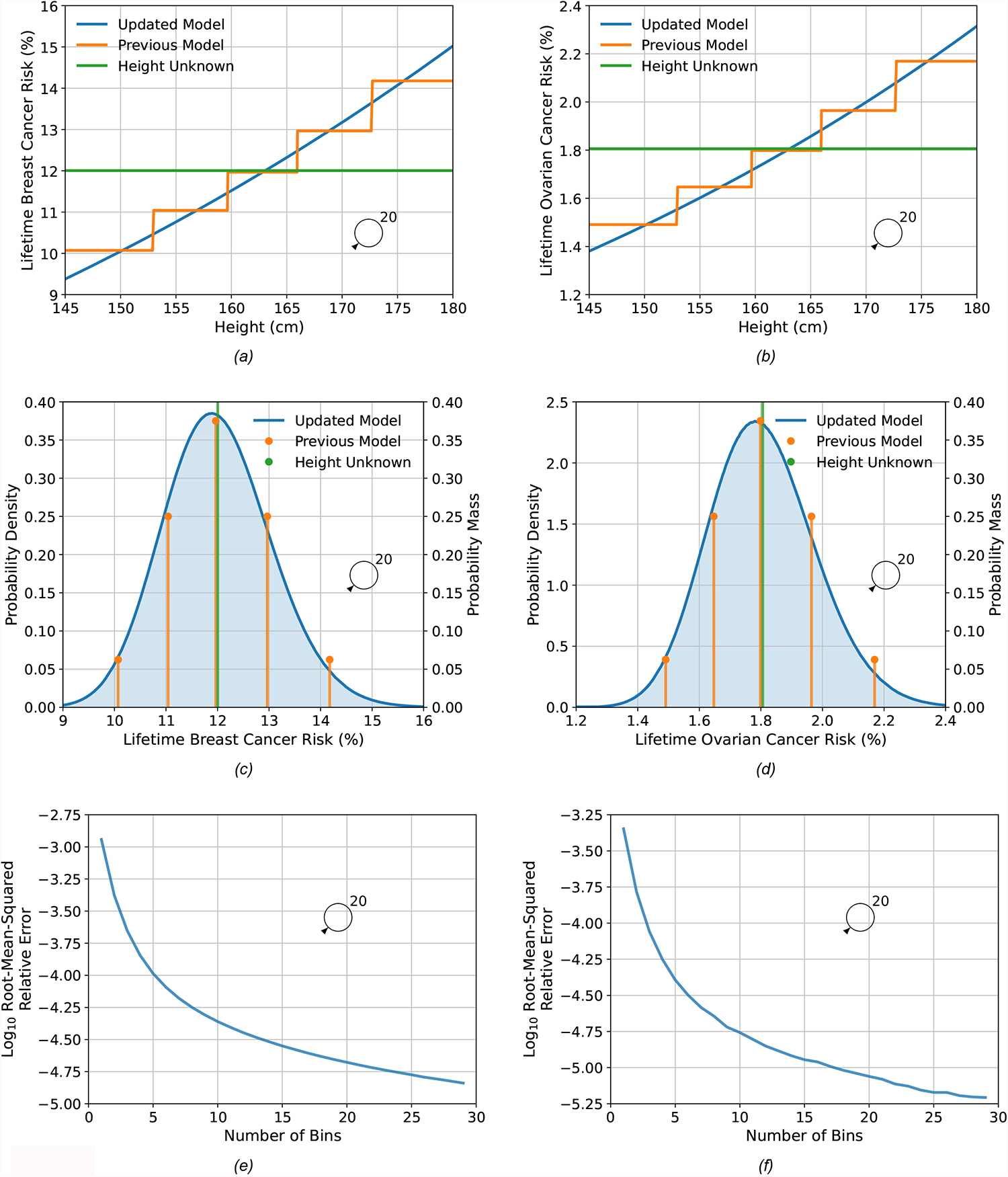
Predicted lifetime breast and ovarian cancer risks as a function of height for a female born in 1985 with unknown family history, comparing the updated model, where height is treated as continuous, to the previous model, where height was treated as categorical. Figures (a), (c) and (e) show breast cancer, while Figures (b), (d) and (f) show ovarian cancer risks. Figures (a) and (b) show the predicted risk as a function of height, while Figures (c) and (d) show the probability density/mass of risk as a function of height. Predictions are based on UK cancer incidences. Figures (e) and (f) show the log (base 10) of the root-mean-squared relative discretisation error as a function of the number of bins. The error was taken to be the absolute difference between the value and the asymptotic extrapolation of the measurements as a function of the number of bins. The average is taken over 100 heights that are spaced 1% apart, from 0.5% to 99.5%.

## Discussion

This work has extended the multifactorial BOADICEA BC and EOC risk prediction models (BOADICEA v6 and the Ovarian Cancer Model v2), employing a synthetic approach^23^. The explicit effects of PVs in *RAD51C, RAD51D, BARD1* and *PALB2*, which are now commonly included on cancer gene panels, are now included in the models. The models have also been extended to accommodate continuous RFs, and parameterisation of tumour pathology and cancer incidences have been updated with more recent data. These represent the most comprehensive models for BC and EOC and will allow more complete BC and EOC risk assessment of those undergoing gene-panel testing.

By explicitly modelling the effects of PVs in the new cancer susceptibility genes, the models provide personalised cancer risks of PV carriers when combined with QRFs, MD and PRS. Although the number of women affected by these changes will be small at population level, for women with *RAD51C, RAD51D* an *BARD1* PVs and their families, the updated risks will be clinically important. *RAD51C, RAD51D* and *BARD1* (like ATM and CHEK2) would be classified as “moderate risk” BC genes based on the average risks^15^. However, according to the BOADICEA predictions, over half (56-59%) of carriers of PVs in these genes would be reclassified from moderate BC risk category to either near-population-risk (34-44%) or high-risk (15-22%), if data on the other risk factors were incorporated (Table s1). Such changes may have important implications for discussions around earlier or more frequent screening or on risk-reduction options for these women. Similarly, based on the multifactorial EOC model, PALB2 PV carriers without EOC family history will always have EOC risks <3% by age 50. However, ∼38% of PALB2 PV carriers will have lifetime EOC risks of >5% (Table s2), which may influence recommendations on the timing of risk-reducing surgery.

As previously, the models assume that the effects of the PVs in the new genes interact multiplicatively with the PRS and the RFs. No studies have yet assessed the joint effects of PVs in these genes and the PRS or RFs. Previous results for *CHEK2* and *ATM* suggest that the multiplicative model holds true for earlier versions of the PRS^31-33^. Unlike *CHEK2* and *ATM*, however, the new genes predispose more strongly to ER-negative disease, and the combined effect may depart from the multiplicative assumption. Demonstrating this explicitly for the new genes will be challenging given the rarity of the mutations. The multiplicative model has also been shown to be reasonable for the combined effects of PRS and RFs^34^, but there is as yet no large-scale evaluation of the combined effects of PVs and RFs. However, recent prospective validation studies of the current and previous versions of the models suggest that, overall, the models fit well^11^[Yang et al. under review]. Should deviations from the multiplicative model between these PVs and RFs emerge, the model can be updated to take them into account.

Both the BC and EOC models incorporate PVs’ effects using the estimated population allele frequencies and RRs. These are combined with reference population incidences to calculate absolute risks while constraining the overall incidences over the RFs included in the model. Our implementation used RR and allele frequency estimates from the largest available studies on those of European ancestry^15^. These were assumed to be constant across all countries. Available data are currently too sparse to obtain country-specific estimates. Although there is no evidence that RRs vary among populations, the allele frequencies are likely to vary to some extent ^15^. This is most apparent for *CHEK2*, where the founder c.1100delC variant is common in northwest Europe and explains the majority of carriers but is rare or absent in other populations. If population-specific variants can be generated, the model can be easily updated to accommodate these. Nevertheless, by allowing population incidences to vary by country, the predicted absolute risks given by the models are country-specific.

The updated age-specific distributions of tumour ER and TN status for six of the BC susceptibility genes in the model (*PALB2, CHEK2, ATM, BARD1, RAD51C* and *RAD51D*) should allow better differentiation between PVs that may be present in a family. Since PV carrier probabilities are used internally in the models, these will also impact the predicted absolute risks for all unaffected individuals if information on tumour characteristics is available for affected relatives whether or not they carry a PV.

We have developed a novel methodological approach for including continuous RFs into the models. We demonstrated this by including height in both the BC and EOC models, allowing for more nuanced predictions and improving the risk discrimination. While the resulting discrimination based on height alone is modest, the framework will allow other more predictive RFs to be included in the model if accurate risk estimates become available. The most important example is MD: continuous measures of MD, available through tools such as STRATUS, CUMULUS and Volpara^35-37^, have been shown to have stronger associations with BC risk than the categorical BI-RADS system. Other examples include BMI and ages at menarche and menopause. Further, the method could be applied to the joint distribution of several continuous risk factors, where the integrals in equations (2) and (3) become multidimensional integrals.

We have further refined the method for creating cohort incidences from calendar period incidences (supplementary material). The approach provides incidences that are less sensitive to year-on-year fluctuations by averaging over all years in the birth cohort. This method is particularly useful for cancers with low incidences, such as EOC and male BC, where the population size is small, and there is no prior averaging over calendar years. The refinement will have little effect on incidences from larger countries.

Our models have certain limitations. No single dataset containing all the required information was available to construct the multifactorial models, so the models were extended via a synthetic approach. The new model parameters were taken from extensive, well-designed published studies together with existing parameters from model fitting^9 10^. We and others have used this approach for developing previous versions of the models^8 11 12 21 38 39^, which have been shown to provide clinically valid predictions^40 41^. Furthermore, the BOADICEA model presented here, has been validated in a large independent study, and has been demonstrated to be well calibrated to discriminate well between affected and unaffected women [Yang et al, under review]. As is the case for the previous versions, the updates presented here are primarily based on studies of those of European ancestry in developed countries. There is little evidence that the RRs associated with PVs differ by ancestry. The PV frequencies are also broadly similar across populations, except for specific founder mutations and CHEK2 PVs, which have a much higher frequency in European than non-European populations. However, other parameters in the model, including RF and PRS distributions, will differ by population, and the model will need to be adapted for use in non-European ancestry populations and developing countries. The synthetic approach presented here allows the model to be easily customised to other populations as better estimates become available^42 43^.

The new model features have been built on the established and well-validated BOADICEA and EOC models^8 11 40^. The updated models will allow for more personalised risks, thus allowing more informed choices on screening, prevention, risk factor modification or other risk-reducing options. The models are available for use by healthcare professionals through the user-friendly CanRisk webtool (www.canrisk.org).

## Data Availability

No individual level data used. The algorithm development was performed using publicly available parameter estimates.
The algorithms developed are freely available for use via www.canrisk.org

## Additional Information

## Authors’ contributions

Conceptualization: ACA, PPDP, DFE

Methodology: AL, ACA, DFE, PPDP

Software: AL, APC, TC, SA, FMW, MT, JR, JUS

Investigation: AL, ACA, PPDP, DFE

Formal analysis: AL, NM

Funding acquisition: ACA, MKS, PD, JS

Resources: PPDP, DFE, VZ, HJ, EF, AdP, MR, TM

Supervision: ACA

Visualisation: AL

Writing – original draft: AL, ACA, DFE

Writing – review & editing: all authors

## Ethics approval and consent to participate

No ethical approval or consent was necessary for this work.

## Data availability

Both models are freely available for non-commercial use through www.canrisk.org

