## Supplementary Material for "Enhancing the BOADICEA cancer risk prediction model to incorporate new data on *RAD51C, RAD51D, BARD1*, updates to tumour pathology and cancer incidences"

#### Pathogenic Variants in Cancer Susceptibility Genes

##### Model definition

The BOADICEA and EOC models assume that the cancer incidences for individual $i$ at age $t$, $\lambda^{(i)}(t)$, depend on their underlying genotype through a model of the form:

$$\lambda^{(i)}(t) = \lambda_0(t)\exp\left(\sum_{\mu=1}^{N_{MG}+1}\left[\left(\beta_{MG\mu}(t) + \sum_{\rho}\boldsymbol{\beta}_{RF\rho\mu}(t)\cdot\boldsymbol{z}_{RF\rho}^{(i)}\right)\prod_{\nu=1}^{\mu-1}\left[\left(1-G_{\nu}^{(i)}\right)G_{\mu}^{(i)}\right]\right] + \beta_{PG}(t)\,x_P^{(i)}\right), \tag{1}$$

where $\lambda_0(t)$ is the baseline incidence (applicable to a non-PV carrier with a zero polygenotype and unknown RFs). $N_{MG}$ is the number of major genes present in the model, which for the previous versions of both models was five. $G_{\mu}^{(i)}$ are indicator variables for the presence/absence of a PV in a major gene in person $i$, taking values 1 if a PV is present and 0 otherwise with $\mu = 1, \ldots, N_{MG}$ representing the genes present in the model in the dominance order and $\mu = N_{MG} + 1$ corresponding to non-carriers of PVs, where $G_{N_{MG}+1}^{(i)} = 1$ for non-carriers of any PV and 0 otherwise. The cancer incidences associated with homozygous and heterozygous carriers of PVs in each gene are assumed to be the same, and the risk to carriers of PVs in more than one gene is assumed to be that of the higher-ranked PV in the dominance order. Because PVs are rare, this model can be well approximated by assuming a single locus with $N_{MG} + 1$ alleles, one representing the presence of a PV in each of the $N_{MG}$ genes and an additional wild-type allele representing absence of PVs in all genes[1]. $\beta_{MG\mu}(t)$ represent the age-specific log-relative risks (log-RRs) associated with the major genes relative to the baseline incidence. The relative risks (RR) assumed for the major genes are summarised in Table 1. $x_P^{(i)}$ is the polygenotype for individual $i$, assumed normally distributed in the general population with mean 0 and standard deviation 1, and $\beta_{PG}(t)$ is the age-specific log-RR per standard deviation associated with the polygene, relative to the baseline incidence[2 3]. When a PRS is known, the polygenotype is decomposed into an observed and residual component where the observed component is given by the PRS[4]. $\rho$ indexes the RFs that are present in the model, which are modelled as categorical factors. $\boldsymbol{\beta}_{RF\rho\mu}(t)$ is the vector (length $\kappa_\rho - 1$ were $\kappa_\rho$ is the number of categories for RF $\rho$, with one category being the baseline) of age-specific log-RRs associated with RF $\rho$, which may depend on the major genotype $\mu$, and $\boldsymbol{z}_{RF\rho}^{(i)}$ is the corresponding vector of indicator variables (0 or 1) that indicate the category of RF $\rho$ for individual $i$ (1 for the observed category, 0 otherwise, with all elements 0 for the baseline). The baseline incidences $\lambda_0(t)$ are determined so that the total age-specific incidences, summed over the RFs and genotypes, agree with the population incidence (given the assumed population distributions and RRs)[2 5]. The population incidences are birth-cohort and country-specific, but this dependence is omitted from equation (1) for clarity of notation. The RRs and distributions of the RF have been described elsewhere[4 6]. To allow appropriately for missing

RF information, only those RFs measured on a given individual are considered (thus, the baseline incidence, $\lambda_0(t)$ are determined for each individual dependent on their measured RFs).

The models assume that RRs associated with PVs in the major genes are log-additive (multiplicative) with the RFs and the polygenic component. The model also assumes that the PVs and the PRS combine multiplicatively (conditional on other factors).

The models evaluate pedigree likelihoods using the MENDEL software[7]. As MENDEL considers only finite discrete genotypes, the polygenotype is approximated by the hypergeometric polygenic model [1 5 8].

Both models consider FH of BC, EOC, pancreatic cancer (PaC) and prostate cancer (PrC). The incidences of each cancer are assumed independent, conditional on the genotypes and RFs in the model. In BOADICEA, EOC, PaC and PrC are assumed to depend only on the major genotype. Correspondingly, in the EOC model, BC, PaC and PrC are assumed to depend only on the major genotype.

#### Adjusting the residual polygenic component after the inclusion of new major genes

The variance due to PVs in each gene at age $t$ is given by:

$$var(t,\mu) = \log\left(\frac{(1-f_\mu)^2 + f_\mu(2-f_\mu)\exp(2\,\beta_{MG\mu}(t))}{\left((1-f_\mu)^2 + f_\mu(2-f_\mu)\exp(\beta_{MG\mu}(t))\right)^2}\right),$$

where $f_\mu$ is the population allele frequency of gene $\mu$; the variance components are assumed to be additive. This process also considered the updated RR and PV frequencies for the previously included genes. For BOADICEA, the overall BC polygenic variance was $4.83 - 0.5961 \times t$ for females and $1.4$ for males, while for the EOC model, the overall EOC polygenic variance was $1.434$ [2 3].

#### Allele Frequencies

Allele frequencies for all genes, except *BRCA1* and *BRCA2,* were taken from the BRIDGES study [9]. The frequencies were based on the frequency of protein-truncating variants in European ancestry controls. To account for the incomplete sensitivity of the sequencing as performed in BRIDGES, the frequencies were adjusted by dividing by $cs(1-v)$, where $c$ is the proportion of the coding sequence of each gene determined to be callable, $s$ is the proportion of variants in the called sequence across all genes that were detected (estimated to be 0.957), and $v$ is the proportion of the pathogenic variants expected to be copy variants. For *CHEK2*, the adjustment was applied to variants excluding c.1100delC. Details are given in the Supplementary Material of Dorling *et al.*[9]. For *BRIP1,* $v$ was assumed to be 0.05. The *BRCA1* and *BRCA2* frequencies from the previous versions of BOADICEA and the EOC model were used for consistency.

##### Sensitivities

The default sensitivities are based on the assumption that protein truncating variants and known pathogenic missense variants are detected with close to 100% sensitivity in clinical tests but that, except for *BRCA1* and *BRCA2*, large rearrangements are not detected. The sensitivities are therefore given by $1 - v$, as above. For *BRCA1* and *BRCA2*, sensitivities were defined by assuming that the main source of insensitivity was missense variants not classified as pathogenic – the frequencies of these variants have been estimated by Dorling et al[10].

#### Population Incidences

The BOADICEA and epithelial tubo-ovarian cancer (EOC) models both allow population customisation via population-specific incidences[4 6 11]. Here the models are extended with incidences from the Netherlands, France, Slovenia and Estonia. Incidences for the Netherlands were taken from Statistics Netherland for 1950-1988 and the Netherlands Cancer Registry for 1989-2017, where BC incidences exclude ductal carcinomas in situ, as these are not included in the models [12 13]. Incidences for France were taken from CI5Plus and CI5 for 1977-1989 using nine registries and from INCa/Santé Public France for 1990-2018 [14-16]. Incidences for Slovenia covering 1961-2016 were taken from the Slovenian Cancer Registry [17]. Incidences for Estonia covering 1968-2018 were taken from the Estonian National Institute for Health Development [18]. Predicted lifetime breast and EOC risks using these incidences are shown in Figure s1.

Incidences for some of the existing regions were updated using data from more recent calendar years. For the UK, incidences covering 2011-2017 were added [19]. For Denmark, Finland, Iceland, Norway and Sweden, incidences covering 2011-2018 were added [20 21]. For Australia, incidences covering 2011-2017 were added [22]. For the USA, incidences covering 2013-2018 from 21 registries were added [23]. For New Zealand, incidences covering 2010-2018 were added [24 25]. For Canada, incidences covering 2011-2018 were added [26]. Figure s2 (a) shows the updated incidences' effects on the cohort incidences for UK female breast cancer incidences for those born in the 1980s.

The models use calendar-specific population incidences to calculate cohort-specific incidences [2], where the cohorts are defined by decadal birth year ranges (1910-1919, 1920-1929, 1930-1939, 1940-1949, 1950-1959, 1960-1969, 1970-1979 and 1980-1989 with individuals born before/after the first/last cohort, assumed to have the same incidences as the first/last cohort). The original model used UK incidences from CI5, which reported calendar incidences averaged in 5-year calendar-period bins [2 14]. Cohort incidences were then taken as those for someone born in the middle year of each range to represent that cohort (1915 for 1910-1919 etc.). However, some of the other regions have smaller populations and report annual-calendar-period specific incidences. For these populations, especially for cancers with low incidences (e.g., EOC and male BC), using a single year to represent the cohort can lead to cohort incidences dominated by year-on-year calendar fluctuations. The methodology was refined by deriving new sets of cohort incidences. In these, the age-specific incidences for an individual in the cohort were taken as the average of the age-specific incidences applicable to those born in each year of the birth-cohort range. The average age- and cohort-specific incidences were then smoothed using LOWESS with linear regression and a bandwidth of 0.2. Figure s2 (b) shows the effects of the new averaging method on cohort incidences for Estonian male breast cancer incidences for those born in the 1920s.

Further, previously, incidences for years before/after the earliest/latest calendar year were taken to be the same as those in the earliest/latest calendar year available. Again, for regions with small populations presenting annual calendar-period incidences and cancers with low incidences, the cohort incidences can be adversely affected by statistical anomalies present in incidences of the earliest/latest calendar year. The methodology was refined with incidence for years before/after the earliest/latest calendar year taken as the average of the first/last five years of the available annual calendar-period incidences.

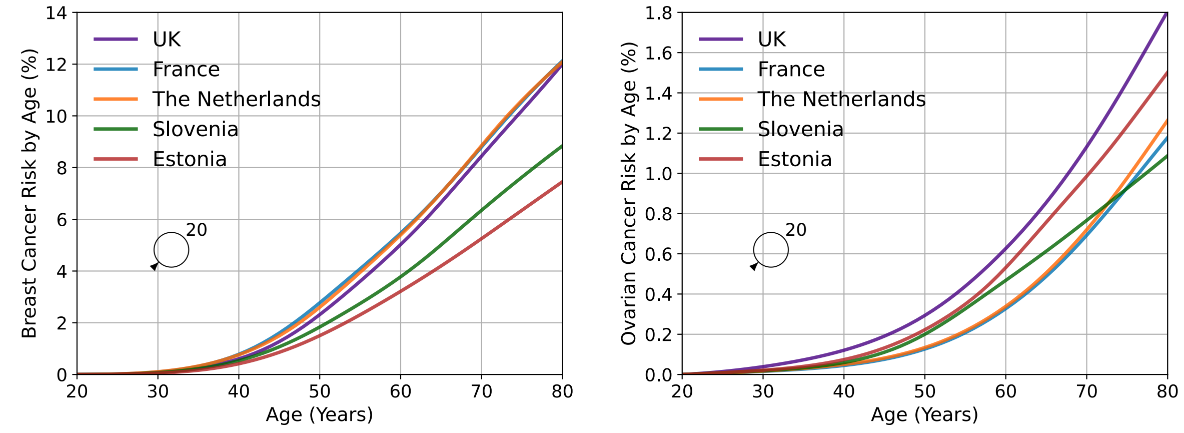


(a) (b)

**Figure s1. Predicted lifetime (age 20 to 80 years) breast and ovarian cancer risk by age for a female born in 1985 with unknown family history (ie average female in the population) comparing risks using incidences for the UK, France, the Netherlands, Slovenia, and Estonia. Figure (a) shows breast cancer risks, where risks for the UK, France, the Netherlands, Slovenia, and Estonia are 12.0%, 12.1%, 12.1%, 8.8% and 7.4%, respectively. Figure (b) shows ovarian cancer risks, where risks for the UK, France, the Netherlands, Slovenia, and Estonia are 1.8%, 1.2%, 1.3%, 1.1% and 1.5%, respectively.**

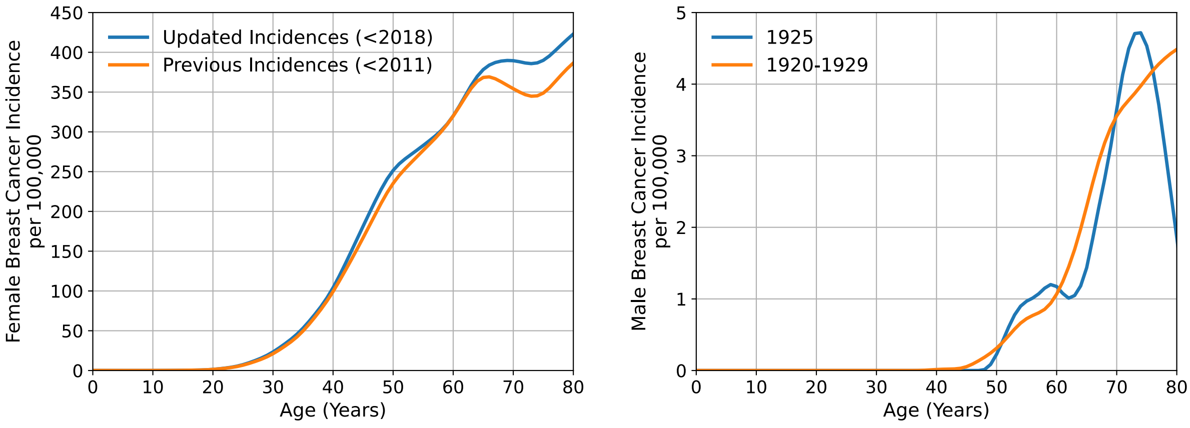


(a) (b)

**Figure s2. Smoothed Cohort-specific population incidences. Figure (a) shows female breast cancer incidences for the UK for those born in the 1980s for the previous incidences (using incidences up to and including 2010) and for the updated incidences (using incidences up to and including 2017), where both datasets use the average over the birth years in the cohort. Figure (b) shows male breast cancer incidences for Estonia for those born in the 1920s, using incidence from a single birth year to represent the cohort (labelled 1925) and using the average over the birth years in the cohort (labelled 1920-1929).**

### New Susceptibility Genes

#### Breast Cancer: *BARD1*, *RAD51C* and *RAD51D*

**Table s1. Predicted 10-year (age 40 to 50 years) and lifetime (age 20 to 80 years) breast cancer risk for a female born in 1985 with unknown family history and for a female with a mother affected at age 50. The columns labelled "Risk" contain risks in the absence of information about questionnaire-based risk factors (QRF), mammographic density (MD) or a polygenic risk score (PRS). The other columns show the distribution of females based on these risk factors in the risk categories defined in the NICE familial breast cancer guidelines [27]: 1) near-population risk, shaded pink ($< 17\%$ lifetime risk; $< 3\%$ 10-year risk), 2) moderate risk, shaded yellow ($\geq 17\%$ and $< 30\%$ lifetime risk; $\geq 3\%$ and $< 8\%$ 10-year risk) and 3) high risk, shaded blue ($\geq 30\%$ lifetime risk; $\geq 8\%$ 10-year risk). Column headings are shaded the same colours as the corresponding lines in Figure 2. Predictions are based on UK cancer incidences.**

| Family History | Risk Horizon | PV Screening | Risk | QRF | | | QRF & MD | | | PRS | | | QRF & PRS | | | QRF, MD & PRS | | |
|---|---|---|---|---|---|---|---|---|---|---|---|---|---|---|---|---|---|---|
| | | | | Pop | Mod | High | Pop | Mod | High | Pop | Mod | High | Pop | Mod | High | Pop | Mod | High |
| Unknown | 10-Year | Untested | 1.7 | 96.4 | 3.6 | 0 | 94.9 | 5.1 | 0 | 90 | 10 | 0 | 90.6 | 9.2 | 0.2 | 89.2 | 10.6 | 0.3 |
| | | *BARD1* | 3.6 | 48.8 | 50.5 | 0.6 | 48.8 | 49.8 | 1.4 | 46.5 | 50 | 3.5 | 56.5 | 39.5 | 4 | 57.8 | 37 | 5.2 |
| | | *RAD51C* | 3.4 | 57.3 | 42.3 | 0.4 | 54 | 44.9 | 1.1 | 50.8 | 46.5 | 2.7 | 60.2 | 36.6 | 3.2 | 61 | 34.7 | 4.4 |
| | | *RAD51D* | 3.1 | 67.3 | 32.5 | 0.2 | 61.1 | 38.2 | 0.7 | 56.5 | 41.6 | 1.8 | 64.9 | 32.8 | 2.3 | 65.2 | 31.4 | 3.4 |
| | | No PV | 1.6 | 97.1 | 2.9 | 0 | 95.8 | 4.2 | 0 | 91.5 | 8.4 | 0 | 91.9 | 8 | 0.1 | 90.5 | 9.3 | 0.2 |
| | Lifetime | Untested | 12 | 92.1 | 7.7 | 0.2 | 88.8 | 10.6 | 0.5 | 87.5 | 12.3 | 0.2 | 85.2 | 13.7 | 1.1 | 83.4 | 14.9 | 1.7 |
| | | *BARD1* | 23.5 | 12.5 | 77.1 | 10.4 | 21.8 | 62.8 | 15.3 | 19.8 | 62.1 | 18.1 | 30.8 | 49.4 | 19.8 | 33.9 | 44.1 | 21.9 |
| | | *RAD51C* | 22.3 | 18.9 | 72.7 | 8.4 | 27.5 | 60.3 | 12.2 | 24.8 | 61 | 14.2 | 35.6 | 47.8 | 16.6 | 38.3 | 42.9 | 18.8 |
| | | *RAD51D* | 20.8 | 29.7 | 63.9 | 6.4 | 35.8 | 55.3 | 8.9 | 32.4 | 57.7 | 9.9 | 42.4 | 44.7 | 12.9 | 44.3 | 40.7 | 15 |
| | | No PV | 11.6 | 92.9 | 7 | 0.1 | 90.1 | 9.4 | 0.5 | 89.1 | 10.7 | 0.1 | 86.7 | 12.4 | 1 | 84.8 | 13.7 | 1.5 |
| Mother affected at age 50 | 10-Year | Untested | 3.5 | 51.8 | 47.6 | 0.5 | 50.6 | 48.1 | 1.3 | 53.7 | 45.7 | 0.6 | 63.4 | 35.1 | 1.5 | 63.5 | 34.1 | 2.4 |
| | | *BARD1* | 6.4 | 1.1 | 87.2 | 11.7 | 10 | 71.9 | 18.1 | 12.5 | 69 | 18.5 | 23.3 | 60.3 | 16.4 | 28.6 | 53.1 | 18.3 |
| | | *RAD51C* | 6.1 | 2 | 88.5 | 9.5 | 12.2 | 73 | 14.8 | 15 | 69.7 | 15.4 | 26.4 | 59.5 | 14.1 | 31.4 | 52.5 | 16.1 |
| | | *RAD51D* | 5.6 | 3.8 | 88.9 | 7.3 | 15.8 | 73 | 11.1 | 18.7 | 69.5 | 11.8 | 30.8 | 57.8 | 11.4 | 35.4 | 51.2 | 13.4 |
| | | No PV | 3.1 | 68.7 | 31.1 | 0.3 | 62.2 | 36.9 | 0.8 | 64.8 | 34.9 | 0.3 | 72 | 26.9 | 1.1 | 71.3 | 26.9 | 1.8 |
| | Lifetime | Untested | 19.3 | 40.6 | 54.6 | 4.8 | 43.5 | 50.1 | 6.3 | 47.2 | 50.3 | 2.5 | 54.6 | 39.2 | 6.2 | 55.1 | 36.8 | 8.1 |
| | | *BARD1* | 33.7 | 0.1 | 37.9 | 62 | 3.4 | 37.4 | 59.3 | 2.3 | 43.2 | 54.6 | 7.8 | 44.2 | 47.9 | 12 | 40.2 | 47.8 |
| | | *RAD51C* | 32.2 | 0.2 | 47.4 | 52.4 | 4.1 | 43.3 | 52.6 | 3.4 | 48.8 | 47.8 | 10 | 47.3 | 42.7 | 14.5 | 42.3 | 43.2 |
| | | *RAD51D* | 30.4 | 0.5 | 59.3 | 40.2 | 5.5 | 50.7 | 43.8 | 5.6 | 55.5 | 38.9 | 13.6 | 50.3 | 36 | 18.3 | 44.4 | 37.3 |
| | | No PV | 17.9 | 55 | 41.3 | 3.8 | 53.5 | 41.6 | 4.9 | 57.8 | 40.5 | 1.7 | 62.6 | 32.5 | 4.9 | 62.1 | 31.3 | 6.6 |

#### Ovarian cancer: *PALB2*

**Table s2. Predicted ovarian cancer risk to age 50 (age 20 to 50 years) and lifetime risk (age 20 to 80 years) for a female born in 1985 with unknown family history and for a female with a mother affected at age 50. The columns labelled "Risk" contain risks in the absence of information about risk factors (RF) or a polygenic risk score (PRS). The other columns show the distribution of females based on these risk factors falling into risk categories defined as: 1) near-population risk, shaded pink ($< 5\%$ lifetime risk; $< 3\%$ risk to age 50), 2) moderate risk, shaded yellow ($\geq 5\%$ and $< 10\%$ lifetime risk; $\geq 3\%$ and $< 5\%$ risk to age 50) and 3) high risk, shaded blue ($\geq 10\%$ lifetime risk; $\geq 5\%$ risk to age 50). Column headings are shaded the same colours as the corresponding lines in Figure 1. Predictions are based on UK cancer incidences.**

| Family History | Risk Horizon | PV Screening | Risk | RF | | | PRS | | | RF & PRS | | |
|---|---|---|---|---|---|---|---|---|---|---|---|---|
| | | | | Pop | Mod | High | Pop | Mod | High | Pop | Mod | High |
| Unknown | Risk to age 50 | Untested | 0.3 | 99.9 | 0 | 0.1 | 100 | 0 | 0 | 100 | 0 | 0 |
| | | *PALB2* | 0.8 | 99.9 | 0 | 0.1 | 100 | 0 | 0 | 100 | 0 | 0 |
| | | No PV | 0.3 | 99.9 | 0 | 0.1 | 100 | 0 | 0 | 100 | 0 | 0 |
| | Lifetime | Untested | 1.8 | 99.9 | 0.1 | 0.1 | 100 | 0 | 0 | 99.6 | 0.4 | 0 |
| | | *PALB2* | 5 | 61.8 | 37.3 | 1 | 55.8 | 44 | 0.2 | 62.4 | 34.9 | 2.7 |
| | | No PV | 1.7 | 99.9 | 0 | 0.1 | 100 | 0 | 0 | 99.7 | 0.3 | 0 |
| Mother affected at age 50 | Risk to age 50 | Untested | 1 | 99.9 | 0 | 0.1 | 100 | 0 | 0 | 100 | 0 | 0 |
| | | *PALB2* | 1.5 | 98.4 | 1.6 | 0.1 | 100 | 0 | 0 | 97.5 | 2.4 | 0.1 |
| | | No PV | 0.5 | 99.9 | 0 | 0.1 | 100 | 0 | 0 | 100 | 0 | 0 |
| | Lifetime | Untested | 5 | 60 | 39.4 | 0.5 | 57.7 | 42.2 | 0 | 62.8 | 36.2 | 1.1 |
| | | *PALB2* | 9.6 | 6 | 58.3 | 35.7 | 0.4 | 65.2 | 34.5 | 11.2 | 55.8 | 33 |
| | | No PV | 3.3 | 91.3 | 8.6 | 0.1 | 98.4 | 1.6 | 0 | 90.1 | 9.7 | 0.1 |

### Tumour Pathology Subtypes

| AGE | GENERAL POPULATION | *BRCA1* | *BRCA2* | *PALB2* | *CHEK2* | *ATM* | *BARD1* | *RAD51C* | *RAD51D* |
|---|---|---|---|---|---|---|---|---|---|
| **20** | 0.4615 | 0.8201 | 0.1881 | 0.5316 | 0.2884 | 0.2366 | 0.7706 | 0.7471 | 0.6255 |
| **21** | 0.4615 | 0.8201 | 0.1881 | 0.5316 | 0.2884 | 0.2366 | 0.7706 | 0.7471 | 0.6255 |
| **22** | 0.4615 | 0.8201 | 0.1881 | 0.5316 | 0.2884 | 0.2366 | 0.7706 | 0.7471 | 0.6255 |
| **23** | 0.4615 | 0.8201 | 0.1881 | 0.5316 | 0.2884 | 0.2366 | 0.7706 | 0.7471 | 0.6255 |
| **24** | 0.4614 | 0.8201 | 0.1881 | 0.5316 | 0.2884 | 0.2366 | 0.7706 | 0.7471 | 0.6255 |
| **25** | 0.4606 | 0.8201 | 0.1881 | 0.5316 | 0.2884 | 0.2366 | 0.7706 | 0.7471 | 0.6255 |
| **26** | 0.459 | 0.8201 | 0.1881 | 0.5218 | 0.2813 | 0.2296 | 0.7633 | 0.7394 | 0.6159 |
| **27** | 0.4567 | 0.8201 | 0.1881 | 0.512 | 0.2743 | 0.2227 | 0.7558 | 0.7314 | 0.6061 |
| **28** | 0.4539 | 0.8201 | 0.1881 | 0.5021 | 0.2674 | 0.216 | 0.748 | 0.7232 | 0.596 |
| **29** | 0.4504 | 0.8201 | 0.1881 | 0.4921 | 0.2605 | 0.2093 | 0.74 | 0.7147 | 0.5858 |
| **30** | 0.4444 | 0.8201 | 0.1881 | 0.482 | 0.2537 | 0.2027 | 0.7318 | 0.706 | 0.5754 |
| **31** | 0.435 | 0.8201 | 0.1881 | 0.4718 | 0.2469 | 0.1961 | 0.7232 | 0.697 | 0.5647 |
| **32** | 0.4228 | 0.8201 | 0.1881 | 0.4615 | 0.2402 | 0.1897 | 0.7144 | 0.6877 | 0.5539 |
| **33** | 0.4093 | 0.8201 | 0.1881 | 0.4513 | 0.2337 | 0.1835 | 0.7055 | 0.6784 | 0.5431 |
| **34** | 0.3953 | 0.8196 | 0.1883 | 0.441 | 0.2272 | 0.1774 | 0.6963 | 0.6688 | 0.5322 |
| **35** | 0.3804 | 0.8171 | 0.1891 | 0.4307 | 0.2207 | 0.1713 | 0.6868 | 0.6588 | 0.521 |
| **36** | 0.3638 | 0.8117 | 0.1908 | 0.4202 | 0.2142 | 0.1653 | 0.677 | 0.6486 | 0.5096 |
| **37** | 0.3461 | 0.804 | 0.1932 | 0.4096 | 0.2079 | 0.1594 | 0.6668 | 0.638 | 0.4979 |
| **38** | 0.3289 | 0.7947 | 0.1962 | 0.3989 | 0.2015 | 0.1536 | 0.6562 | 0.627 | 0.4859 |
| **39** | 0.3139 | 0.7849 | 0.1993 | 0.3881 | 0.1951 | 0.1478 | 0.6451 | 0.6156 | 0.4737 |
| **40** | 0.3013 | 0.775 | 0.2024 | 0.3771 | 0.1888 | 0.142 | 0.6336 | 0.6038 | 0.4611 |
| **41** | 0.2899 | 0.7652 | 0.2055 | 0.3661 | 0.1826 | 0.1364 | 0.6217 | 0.5915 | 0.4483 |
| **42** | 0.2785 | 0.7559 | 0.2085 | 0.3549 | 0.1763 | 0.1308 | 0.6093 | 0.5788 | 0.4353 |
| **43** | 0.2676 | 0.7482 | 0.2109 | 0.3469 | 0.1717 | 0.1269 | 0.6008 | 0.5703 | 0.4266 |
| **44** | 0.2583 | 0.7426 | 0.213 | 0.3389 | 0.1671 | 0.123 | 0.5922 | 0.5615 | 0.4179 |
| **45** | 0.2511 | 0.7388 | 0.2157 | 0.3309 | 0.1625 | 0.1191 | 0.5834 | 0.5525 | 0.409 |
| **46** | 0.2451 | 0.7357 | 0.2197 | 0.3227 | 0.1579 | 0.1153 | 0.5743 | 0.5433 | 0.4001 |
| **47** | 0.2395 | 0.7318 | 0.2253 | 0.3146 | 0.1533 | 0.1115 | 0.565 | 0.5338 | 0.391 |
| **48** | 0.2343 | 0.7273 | 0.232 | 0.3094 | 0.1503 | 0.1092 | 0.5597 | 0.5285 | 0.386 |
| **49** | 0.2301 | 0.7224 | 0.2392 | 0.3043 | 0.1473 | 0.1069 | 0.5544 | 0.523 | 0.3809 |
| **50** | 0.2269 | 0.7176 | 0.2463 | 0.2991 | 0.1443 | 0.1045 | 0.549 | 0.5175 | 0.3759 |
| **51** | 0.2243 | 0.7127 | 0.2535 | 0.294 | 0.1413 | 0.1022 | 0.5435 | 0.512 | 0.3708 |
| **52** | 0.2217 | 0.7082 | 0.2602 | 0.2888 | 0.1383 | 0.0999 | 0.538 | 0.5064 | 0.3657 |
| **53** | 0.2192 | 0.7043 | 0.2658 | 0.2852 | 0.1363 | 0.0984 | 0.5344 | 0.5027 | 0.3624 |
| **54** | 0.2167 | 0.7008 | 0.2697 | 0.2817 | 0.1343 | 0.0968 | 0.5307 | 0.499 | 0.3592 |
| **55** | 0.2139 | 0.695 | 0.2717 | 0.2781 | 0.1323 | 0.0953 | 0.5271 | 0.4953 | 0.3559 |
| **56** | 0.2103 | 0.6852 | 0.2725 | 0.2745 | 0.1303 | 0.0937 | 0.5234 | 0.4916 | 0.3526 |
| **57** | 0.2058 | 0.6713 | 0.273 | 0.2709 | 0.1283 | 0.0922 | 0.5197 | 0.4878 | 0.3493 |
| **58** | 0.2009 | 0.6547 | 0.2736 | 0.2678 | 0.1266 | 0.0908 | 0.5164 | 0.4845 | 0.3464 |
| **59** | 0.1961 | 0.6371 | 0.2743 | 0.2646 | 0.1249 | 0.0895 | 0.5131 | 0.4812 | 0.3435 |
| **60** | 0.1911 | 0.6193 | 0.2749 | 0.2614 | 0.1232 | 0.0882 | 0.5098 | 0.4778 | 0.3407 |
| **61** | 0.1858 | 0.6017 | 0.2756 | 0.2583 | 0.1214 | 0.0868 | 0.5064 | 0.4745 | 0.3378 |
| **62** | 0.1803 | 0.5851 | 0.2762 | 0.2551 | 0.1197 | 0.0855 | 0.5031 | 0.4711 | 0.3349 |
| **63** | 0.1751 | 0.5712 | 0.2767 | 0.2524 | 0.1185 | 0.0844 | 0.5 | 0.468 | 0.3322 |
| **64** | 0.1707 | 0.5617 | 0.2771 | 0.2498 | 0.1172 | 0.0833 | 0.4969 | 0.4649 | 0.3295 |
| **65** | 0.1677 | 0.5571 | 0.2773 | 0.2471 | 0.1159 | 0.0823 | 0.4938 | 0.4618 | 0.3268 |
| **66** | 0.1656 | 0.5562 | 0.2773 | 0.2445 | 0.1146 | 0.0812 | 0.4907 | 0.4587 | 0.3242 |
| **67** | 0.1641 | 0.5562 | 0.2773 | 0.2418 | 0.1133 | 0.0801 | 0.4876 | 0.4556 | 0.3215 |
| **68** | 0.1631 | 0.5562 | 0.2773 | 0.2392 | 0.1121 | 0.079 | 0.4845 | 0.4524 | 0.3188 |
| **69** | 0.1625 | 0.5562 | 0.2773 | 0.2366 | 0.1108 | 0.078 | 0.4813 | 0.4493 | 0.3161 |
| **70** | 0.162 | 0.5562 | 0.2773 | 0.234 | 0.1096 | 0.077 | 0.4782 | 0.4461 | 0.3135 |
| **71** | 0.1613 | 0.5562 | 0.2773 | 0.2314 | 0.1083 | 0.0759 | 0.475 | 0.443 | 0.3108 |
| **72** | 0.1601 | 0.5562 | 0.2773 | 0.2287 | 0.1071 | 0.0749 | 0.4718 | 0.4398 | 0.3081 |
| **73** | 0.1584 | 0.5562 | 0.2773 | 0.2263 | 0.106 | 0.0739 | 0.4688 | 0.4368 | 0.3056 |
| **74** | 0.1564 | 0.5562 | 0.2773 | 0.2239 | 0.1049 | 0.073 | 0.4658 | 0.4338 | 0.3031 |
| **75** | 0.1538 | 0.5562 | 0.2773 | 0.2215 | 0.1038 | 0.072 | 0.4628 | 0.4308 | 0.3006 |
| **76** | 0.1507 | 0.5562 | 0.2773 | 0.219 | 0.1026 | 0.0711 | 0.4597 | 0.4278 | 0.2981 |
| **77** | 0.1474 | 0.5562 | 0.2773 | 0.2166 | 0.1015 | 0.0701 | 0.4566 | 0.4248 | 0.2955 |
| **78** | 0.1446 | 0.5562 | 0.2773 | 0.2142 | 0.1004 | 0.0692 | 0.4536 | 0.4217 | 0.293 |
| **79** | 0.1434 | 0.5562 | 0.2773 | 0.2117 | 0.0993 | 0.0683 | 0.4505 | 0.4186 | 0.2905 |

**Table s3: Age-specific proportion of oestrogen receptor-negative tumours among all female breast cancer tumours in the general population and carriers of pathogenic variants in the breast cancer susceptibility genes used in the BOADICEA model.**

| AGE | GENERAL POPULATION | *BRCA1* | *BRCA2* | *PALB2* | *CHEK2* | *ATM* | *BARD1* | *RAD51C* | *RAD51D* |
|---|---|---|---|---|---|---|---|---|---|
| **20** | 0.6582 | 0.8799 | 0.7586 | 0.6066 | 0.3768 | 0.5979 | 0.8195 | 0.8094 | 0.8489 |
| **21** | 0.6582 | 0.8799 | 0.7586 | 0.6066 | 0.3768 | 0.5979 | 0.8195 | 0.8094 | 0.8489 |
| **22** | 0.6582 | 0.8799 | 0.7586 | 0.6066 | 0.3768 | 0.5979 | 0.8195 | 0.8094 | 0.8489 |
| **23** | 0.6582 | 0.8799 | 0.7586 | 0.6066 | 0.3768 | 0.5979 | 0.8195 | 0.8094 | 0.8489 |
| **24** | 0.6579 | 0.8799 | 0.7586 | 0.6066 | 0.3768 | 0.5979 | 0.8195 | 0.8094 | 0.8489 |
| **25** | 0.6566 | 0.8799 | 0.7586 | 0.6066 | 0.3768 | 0.5979 | 0.8195 | 0.8094 | 0.8489 |
| **26** | 0.6539 | 0.8799 | 0.7586 | 0.6047 | 0.3766 | 0.596 | 0.8183 | 0.8082 | 0.8479 |
| **27** | 0.6499 | 0.8799 | 0.7586 | 0.6027 | 0.3763 | 0.594 | 0.8171 | 0.8069 | 0.8468 |
| **28** | 0.6451 | 0.8799 | 0.7586 | 0.6006 | 0.3759 | 0.5919 | 0.8158 | 0.8055 | 0.8457 |
| **29** | 0.6401 | 0.8799 | 0.7586 | 0.5984 | 0.3754 | 0.5897 | 0.8144 | 0.8041 | 0.8445 |
| **30** | 0.6349 | 0.8799 | 0.7586 | 0.5961 | 0.3748 | 0.5874 | 0.813 | 0.8026 | 0.8433 |
| **31** | 0.6299 | 0.8799 | 0.7586 | 0.5937 | 0.3741 | 0.5849 | 0.8114 | 0.801 | 0.8419 |
| **32** | 0.6251 | 0.8799 | 0.7586 | 0.5911 | 0.3732 | 0.5824 | 0.8098 | 0.7993 | 0.8405 |
| **33** | 0.6211 | 0.8799 | 0.7586 | 0.5888 | 0.3726 | 0.58 | 0.8083 | 0.7977 | 0.8392 |
| **34** | 0.6181 | 0.8799 | 0.7586 | 0.5863 | 0.3719 | 0.5775 | 0.8067 | 0.7961 | 0.8378 |
| **35** | 0.6156 | 0.8799 | 0.7586 | 0.5837 | 0.371 | 0.5749 | 0.805 | 0.7943 | 0.8364 |
| **36** | 0.6128 | 0.8799 | 0.7586 | 0.5809 | 0.3699 | 0.5721 | 0.8032 | 0.7925 | 0.8348 |
| **37** | 0.6091 | 0.8799 | 0.7586 | 0.578 | 0.3688 | 0.5691 | 0.8013 | 0.7905 | 0.8331 |
| **38** | 0.6046 | 0.8799 | 0.7586 | 0.5747 | 0.3673 | 0.5658 | 0.7992 | 0.7882 | 0.8312 |
| **39** | 0.5999 | 0.8799 | 0.7586 | 0.5712 | 0.3655 | 0.5623 | 0.7968 | 0.7858 | 0.8292 |
| **40** | 0.5952 | 0.8799 | 0.7586 | 0.5674 | 0.3636 | 0.5585 | 0.7943 | 0.7832 | 0.827 |
| **41** | 0.5905 | 0.8799 | 0.7586 | 0.5634 | 0.3614 | 0.5545 | 0.7917 | 0.7804 | 0.8247 |
| **42** | 0.586 | 0.8799 | 0.7586 | 0.5591 | 0.359 | 0.5501 | 0.7888 | 0.7774 | 0.8221 |
| **43** | 0.5823 | 0.8799 | 0.7586 | 0.5588 | 0.3604 | 0.5499 | 0.7886 | 0.7772 | 0.822 |
| **44** | 0.5798 | 0.8799 | 0.7586 | 0.5585 | 0.3617 | 0.5496 | 0.7884 | 0.777 | 0.8218 |
| **45** | 0.5785 | 0.8799 | 0.7586 | 0.5583 | 0.3631 | 0.5493 | 0.7882 | 0.7768 | 0.8216 |
| **46** | 0.5783 | 0.8799 | 0.7586 | 0.558 | 0.3644 | 0.549 | 0.788 | 0.7766 | 0.8215 |
| **47** | 0.5783 | 0.8799 | 0.7586 | 0.5576 | 0.3657 | 0.5487 | 0.7878 | 0.7764 | 0.8213 |
| **48** | 0.5783 | 0.8799 | 0.7586 | 0.5602 | 0.3698 | 0.5513 | 0.7895 | 0.7782 | 0.8228 |
| **49** | 0.5783 | 0.8799 | 0.7586 | 0.563 | 0.374 | 0.554 | 0.7914 | 0.7801 | 0.8244 |
| **50** | 0.5783 | 0.8799 | 0.7586 | 0.5658 | 0.3784 | 0.5569 | 0.7933 | 0.7821 | 0.8261 |
| **51** | 0.5783 | 0.8799 | 0.7586 | 0.5687 | 0.3828 | 0.5598 | 0.7952 | 0.7841 | 0.8278 |
| **52** | 0.5783 | 0.8799 | 0.7586 | 0.5718 | 0.3875 | 0.5629 | 0.7972 | 0.7862 | 0.8295 |
| **53** | 0.5783 | 0.8799 | 0.7586 | 0.5749 | 0.3922 | 0.566 | 0.7993 | 0.7884 | 0.8313 |
| **54** | 0.5783 | 0.8799 | 0.7586 | 0.5781 | 0.397 | 0.5692 | 0.8014 | 0.7905 | 0.8332 |
| **55** | 0.5783 | 0.8799 | 0.7586 | 0.5814 | 0.4019 | 0.5726 | 0.8035 | 0.7928 | 0.8351 |
| **56** | 0.5783 | 0.8799 | 0.7586 | 0.5848 | 0.407 | 0.576 | 0.8057 | 0.7951 | 0.837 |
| **57** | 0.5783 | 0.8799 | 0.7586 | 0.5883 | 0.4121 | 0.5795 | 0.808 | 0.7974 | 0.8389 |
| **58** | 0.5783 | 0.8799 | 0.7586 | 0.5915 | 0.4171 | 0.5827 | 0.81 | 0.7996 | 0.8407 |
| **59** | 0.5783 | 0.8799 | 0.7586 | 0.5948 | 0.4221 | 0.5861 | 0.8121 | 0.8018 | 0.8426 |
| **60** | 0.5783 | 0.8799 | 0.7586 | 0.5982 | 0.4273 | 0.5895 | 0.8143 | 0.804 | 0.8444 |
| **61** | 0.5783 | 0.8799 | 0.7586 | 0.6017 | 0.4326 | 0.593 | 0.8165 | 0.8063 | 0.8463 |
| **62** | 0.5783 | 0.8799 | 0.7586 | 0.6053 | 0.438 | 0.5966 | 0.8187 | 0.8086 | 0.8483 |
| **63** | 0.5783 | 0.8799 | 0.7586 | 0.6073 | 0.4417 | 0.5986 | 0.8199 | 0.8099 | 0.8493 |
| **64** | 0.5783 | 0.8799 | 0.7586 | 0.6093 | 0.4455 | 0.6006 | 0.8212 | 0.8111 | 0.8504 |
| **65** | 0.5783 | 0.8799 | 0.7586 | 0.6113 | 0.4494 | 0.6027 | 0.8224 | 0.8125 | 0.8515 |
| **66** | 0.5783 | 0.8799 | 0.7586 | 0.6134 | 0.4533 | 0.6048 | 0.8237 | 0.8138 | 0.8526 |
| **67** | 0.5783 | 0.8799 | 0.7586 | 0.6155 | 0.4572 | 0.6069 | 0.825 | 0.8152 | 0.8537 |
| **68** | 0.5783 | 0.8799 | 0.7586 | 0.6176 | 0.4611 | 0.609 | 0.8262 | 0.8164 | 0.8548 |
| **69** | 0.5783 | 0.8799 | 0.7586 | 0.6196 | 0.4651 | 0.6111 | 0.8275 | 0.8178 | 0.8559 |
| **70** | 0.5783 | 0.8799 | 0.7586 | 0.6218 | 0.4691 | 0.6132 | 0.8288 | 0.8191 | 0.857 |
| **71** | 0.5783 | 0.8799 | 0.7586 | 0.624 | 0.4731 | 0.6154 | 0.8301 | 0.8205 | 0.8581 |
| **72** | 0.5783 | 0.8799 | 0.7586 | 0.6262 | 0.4773 | 0.6177 | 0.8315 | 0.8219 | 0.8593 |
| **73** | 0.5783 | 0.8799 | 0.7586 | 0.6283 | 0.4812 | 0.6197 | 0.8327 | 0.8232 | 0.8603 |
| **74** | 0.5783 | 0.8799 | 0.7586 | 0.6303 | 0.4851 | 0.6218 | 0.8339 | 0.8245 | 0.8614 |
| **75** | 0.5783 | 0.8799 | 0.7586 | 0.6324 | 0.4892 | 0.624 | 0.8352 | 0.8258 | 0.8625 |
| **76** | 0.5783 | 0.8799 | 0.7586 | 0.6346 | 0.4933 | 0.6262 | 0.8365 | 0.8271 | 0.8636 |
| **77** | 0.5783 | 0.8799 | 0.7586 | 0.6368 | 0.4974 | 0.6284 | 0.8378 | 0.8285 | 0.8647 |
| **78** | 0.5783 | 0.8799 | 0.7586 | 0.6391 | 0.5016 | 0.6307 | 0.8391 | 0.8299 | 0.8659 |
| **79** | 0.5783 | 0.8799 | 0.7586 | 0.6415 | 0.5059 | 0.6331 | 0.8405 | 0.8313 | 0.8671 |

**Table s4: Age-specific proportion of triple-negative tumours among female oestrogen receptor-negative breast cancer tumours in the general population and carriers of pathogenic variants in the breast cancer susceptibility genes used in the BOADICEA model.**

Hall P, Hamann U, Harkness EF, Hartikainen JM, Hartman M, He W, Heemskerk-Gerritsen BAM, Hillemanns P, Hogervorst FBL, Hollestelle A, Ho WK, Hooning MJ, Howell A, Humphreys K, Idris F, Jakubowska A, Jung A, Kapoor PM, Kerin MJ, Khusnutdinova E, Kim SW, Ko YD, Kosma VM, Kristensen VN, Kyriacou K, Lakeman IMM, Lee JW, Lee MH, Li J, Lindblom A, Lo WY, Loizidou MA, Lophatananon A, Lubiński J, MacInnis RJ, Madsen MJ, Mannermaa A, Manoochehri M, Manoukian S, Margolin S, Martinez ME, Maurer T, Mavroudis D, McLean C, Meindl A, Mensenkamp AR, Michailidou K, Miller N, Mohd Taib NA, Muir K, Mulligan AM, Nevanlinna H, Newman WG, Nordestgaard BG, Ng PS, Oosterwijk JC, Park SK, Park-Simon TW, Perez JIA, Peterlongo P, Porteous DJ, Prajzendanc K, Prokofyeva D, Radice P, Rashid MU, Rhenius V, Rookus MA, Rüdiger T, Saloustros E, Sawyer EJ, Schmutzler RK, Schneeweiss A, Schürmann P, Shah M, Sohn C, Southey MC, Surowy H, Suvanto M, Thanasitthichai S, Tomlinson I, Torres D, Truong T, Tzardi M, Valova Y, van Asperen CJ, Van Dam RM, van den Ouweland AMW, van der Kolk LE, van Veen EM, Wendt C, Williams JA, Yang XR, Yoon SY, Zamora MP, Evans DG, de la Hoya M, Simard J, Antoniou AC, Borg Å, Andrulis IL, Chang-Claude J, García-Closas M, Chenevix-Trench G, Milne RL, Pharoah PDP, Schmidt MK, Spurdle AB, Vreeswijk MPG, Benitez J, Dunning AM, Kvist A, Teo SH, Devilee P, Easton DF, Consortium BCA. Breast Cancer Risk Genes - Association Analysis in More than 113,000 Women. *N Engl J Med* 2021;384(5):428-39. doi: 10.1056/NEJMoa1913948 [published Online First: 2021/01/20]

10. Dorling L, Carvalho S, Allen J, Parsons MT, Fortuno C, González-Neira A, Heijl SM, Adank MA, Ahearn TU, Andrulis IL, Auvinen P, Becher H, Beckmann MW, Behrens S, Bermisheva M, Bogdanova NV, Bojesen SE, Bolla MK, Bremer M, Briceno I, Camp NJ, Campbell A, Castelao JE, Chang-Claude J, Chanock SJ, Chenevix-Trench G, Collaborators N, Collée JM, Czene K, Dennis J, Dörk T, Eriksson M, Evans DG, Fasching PA, Figueroa J, Flyger H, Gabrielson M, Gago-Dominguez M, García-Closas M, Giles GG, Glendon G, Guénel P, Gündert M, Hadjisavvas A, Hahnen E, Hall P, Hamann U, Harkness EF, Hartman M, Hogervorst FBL, Hollestelle A, Hoppe R, Howell A, Investigators k, Investigators S, Jakubowska A, Jung A, Khusnutdinova E, Kim S-W, Ko Y-D, Kristensen VN, Lakeman IMM, Li J, Lindblom A, Loizidou MA, Lophatananon A, Lubiński J, Luccarini C, Madsen MJ, Mannermaa A, Manoochehri M, Margolin S, Mavroudis D, Milne RL, Mohd Taib NA, Muir K, Nevanlinna H, Newman WG, Oosterwijk JC, Park SK, Peterlongo P, Radice P, Saloustros E, Sawyer EJ, Schmutzler RK, Shah M, Sim X, Southey MC, Surowy H, Suvanto M, Tomlinson I, Torres D, Truong T, van Asperen CJ, Waltes R, Wang Q, Yang XR, Pharoah PDP, Schmidt MK, Benitez J, Vroling B, Dunning AM, Teo SH, Kvist A, de la Hoya M, Devilee P, Spurdle AB, Vreeswijk MPG, Easton DF. Breast cancer risks associated with missense variants in breast cancer susceptibility genes. *medRxiv* 2021:2021.09.02.21262369. doi: 10.1101/2021.09.02.21262369
11. Lee AJ, Cunningham AP, Kuchenbaecker KB, Mavaddat N, Easton DF, Antoniou AC, Consortium of Investigators of Modifiers of B, Breast Cancer Association C. BOADICEA breast cancer risk prediction model: updates to cancer incidences, tumour pathology and web interface. *Br J Cancer* 2014;110(2):535-45. doi: 10.1038/bjc.2013.730
12. Statistics Netherlands. [Available from: www.cbs.nl.
13. Netherlands Cancer Registry. [Available from: www.iknl.nl.

14. Ferlay J, Parkin DM, Curado MP, Bray F, Edwards B, Shin HR, Forman D. Cancer Incidence in Five Continents, Volumes I to IX: IARC CancerBase No. 9 [Internet]: International Agency for Research on Cancer, 2010.
15. Ferlay J, Bray F, Steliarova-Foucher E, Forman D. Cancer Incidence in Five Continents, CI5plus. IARC CancerBase No. 9: Lyon: International Agency for Research on Cancer, 2014.
16. Defossez G, Le Guyader-Peyrou S, Uhry Z, Grosclaude P, Colonna M, Dantony E, Delafosse P, Molinié F, Woronoff A-S, Bouvier A-M, Bossard N, Remontet L, Monnereau A. Estimations nationales de l'incidence et de la mortalité par cancer en France métropolitaine entre 1990 et 2018 - Volume 1: Tumeurs solides : Étude à partir des registres des cancers du réseau Francim. *Santé publique France* 2019:372.
17. Slovenian Cancer Registry. [Available from: https://www.onko-i.si.
18. The Estonian National Institute for Health Development. Age-specific incidence rate of malignant neoplasms per 100 000 inhabitants by site and sex. [updated [Last Update: 2020 September 12]. Available from: https://statistika.tai.ee/pxweb/en/Andmebaas/Andmebaas 02Haigestumus 04PahaloomulisedKasvajad/PK30.px/.
19. Office for National Statistics UK. Cancer Statistics Registrations, England, Series MB1 2017 [updated [Last Update: 2019-05-16]. Available from: https://www.ons.gov.uk/peoplepopulationandcommunity/healthandsocialcare/conditionsanddiseases/datasets/cancerregistrationstatisticscancerregistrationstatisticsengland accessed August 23 2021.
20. Danckert B, Ferlay J, Engholm G, Hansen H, Johannesen T, Khan S, Køtlum J, Ólafsdóttir E, Schmidt L, Virtanen A, Storm H. NORDCAN: Cancer Incidence, Mortality, Prevalence and Survival in the Nordic Countries, Version 8.2 (26.03.2019). Association of the Nordic Cancer Registries. Danish Cancer Society.; 2019 [Available from: http://www.ancr.nu accessed August 18 2021.
21. Engholm G, Ferlay J, Christensen N, Bray F, Gjerstorff ML, Klint A, Kotlum JE, Olafsdottir E, Pukkala E, Storm HH. NORDCAN--a Nordic tool for cancer information, planning, quality control and research. *Acta Oncol* 2010;49(5):725-36. doi: 10.3109/02841861003782017 [doi]
22. Australian Institute of Health and Welfare (AIHW). Australian Cancer Incidence and Mortality (ACIM) Books, CAN 122: AIHW: Canberra; 2021 [updated 08 Jun 2021. accessed August 22 2021.
23. Surveillance Epidemiology and End Results (SEER) Program. SEER*Stat Database: Incidence - SEER 21 Regs Research Data, Nov 2020 - Linked To County Attributes - Total U.S. 2020 [updated 2020. Available from: www.seer.cancer.gov accessed August 18 2021.
24. Lewis C. Personal Communication: New Zealand Cancer Incidences, 2021.
25. New Zealand Ministry of Health. Cancer Statistis. In: Health Mo, ed., 2021.
26. Statistics Canada. Table 13-10-0111-01 Number and rates of new cases of primary cancer, by cancer type, age group and sex 2021 [updated [Last Update: 2020-10-23]. Available from: https://www150.statcan.gc.ca/t1/tbl1/en/tv.action?pid=1310011101 accessed August 22 2021.
27. National Institute for Health and Care Excellence. Familial breast cancer: classification and care of people at risk of familial breast cancer and management of breast cancer and

related risks in people with a family history of breast cancer (CG164) 2013 [updated 2019-11-20. Available from: http://www.nice.org.uk/CG164.
